# A comparison of the genes and genesets identified by EWAS and GWAS of fourteen complex traits

**DOI:** 10.1101/2022.03.25.22272928

**Authors:** Thomas Battram, Tom R. Gaunt, Caroline L. Relton, Nicholas J. Timpson, Gibran Hemani

**Affiliations:** MRC Integrative Epidemiology Unit, University of Bristol, UK; Population Health Sciences, Bristol Medical School, University of Bristol, UK

## Abstract

Identifying the genes, properties of these genes and pathways to understand the underlying biology of complex traits responsible for differential health states in the population is a common goal of epigenome-wide and genome-wide association studies (EWAS and GWAS). GWAS identify genetic variants that effect the trait of interest or variants that are in linkage disequilibrium with the true causal variants. EWAS identify variation in DNA methylation, a complex molecular phenotype, associated with the trait of interest. Therefore, while GWAS in principle will only detect variants within or near causal genes, EWAS can also detect genes that confound the association between a trait and a DNA methylation site, or are reverse causal. Here we systematically compare association EWAS and GWAS results of 14 complex traits (N > 4500). A small fraction of detected genomic regions were shared by both EWAS and GWAS (0-9%). We evaluated if the genes or gene ontology terms flagged by GWAS and EWAS overlapped, and after a multiple testing correction, found substantial overlap for diastolic blood pressure (gene overlap P = 5.2×10^−6^, term overlap P = 0.001). We superimposed our empirical findings against simulated models of varying genetic and epigenetic architectures and observed that in a majority of cases EWAS and GWAS are likely capturing distinct genesets, implying that genes identified by EWAS are not generally causally upstream of the trait. Overall our results indicate that EWAS and GWAS are capturing different aspects of the biology of complex traits.

## Introduction

Determining molecular associations with complex traits can yield novel therapeutic interventions. Large efforts have been put in place to conduct hypothesis free searches to identify associations between disease variation and variation in molecular features. Two popular study designs include genome-wide association studies (GWAS) and epigenome-wide association studies (EWAS). EWAS and GWAS have different statistical properties which may lead them to prioritise different aspects of disease biology. In this paper we investigate the relationship between the results of these two approaches.

EWAS assess the association between one facet of epigenetics, DNA methylation, and complex traits (1–3). Often in EWAS, the potential biological implications of differentially methylated positions or regions (DMPs or DMRs) will be investigated further through genomic annotations (4–7). Previous studies have demonstrated a relationship between DNA methylation levels and proximal genes (8,9). This observation has lead to it being common place to map sites identified in EWAS to nearby genes and these genes and their function are often probed to ascertain their relevance to the trait of interest (4–7). Further, genes can be grouped with others into “genesets” that have similar functionality or lie within the same biological pathway. Examining over-represented genesets may provide an insight into the molecular biology of a trait. For example, Reese et al. performed an EWAS of asthma and discovered the DMPs and DMRs identified mapped to genes within relevant immunological pathways more than expected by chance (5). These genesets included endothelial nitric oxide synthase (eNOS) signaling, the inflammasome, and nuclear factor *κ*B (NF-*κ*B) signaling (5). Other assessments may be made to attempt to infer biological understanding, including enrichment of other epigenetic marks at the regions identified (10) and follow-up experimental studies (1,11). However, using open access databases to investigate tagged genes and genesets is a simple and potentially effective approach to further biological understanding.

GWAS often use similar approaches to EWAS to help discovery of genomic regions related to complex traits (1,12). Examples of genetic-epigenetic equivalence are known, that is the identical, clinically measurable, effects of a genetic lesion and an epigenetic change. For example, Angelman syndrome and Prader Willi syndrome can both be caused by a deletion mutation or by imprinting disorders (13,14). Hence, there is some precedent to expect overlap in biological interpretation between GWAS and EWAS hits. However, the properties of genetic variants and DNA methylation differ in terms of the causal mechanisms that can give rise to trait associations, making potential inferences from EWAS and GWAS diverge. Importantly, DNA methylation is responsive to environmental stimuli, thus making associations identified in EWAS potentially attributable to reverse causation and to confounding (2,3,15). Recent work has suggested that there is a general trend for DNA methylation associations with complex traits being more likely due to confounding or reverse causation than DNA methylation itself being causal (16,17). It should be noted that GWAS may be susceptible to confounding due to various population phenomena such as population stratification or dynastic effects (18), but statistical adjustments are routinely applied to address this, and confounding is unlikely to be affecting GWAS signals, especially for clinical traits (19,20).

A direct comparison between EWAS and GWAS results could provide insight into what biological information EWAS are capturing. If EWAS are highlighting a similar set of genes and genesets to GWAS, it suggests changes in DNA methylation are either themselves involved in trait aetiology or tagging molecular aetiologically relevant genomic features. Similarly, DMPs identified as the result of confounding and reverse causation may also highlight similar genes and genesets as GWAS if these confounding and reverse causal pathways are similar to the causal pathways for a trait. Regardless, in that scenario EWAS is still identifying facets of trait aetiology. In the event that GWAS and EWAS are not highlighting a similar set of genes and genesets, it is plausible that EWAS may still be identifying facets of trait aetiology. Under the circumstance that biological insights from EWAS and GWAS overlap, the efficiency of EWAS would far outstrip that of GWAS, for example an EWAS of BMI using around 10,000 samples identified 187 independent genomic loci (17) while it required a GWAS of 330,000 samples to identify 97 independent genomic loci (21).

If DNA methylation mediates non-genetic effects or if sites are mapped to genes or genesets incorrectly then overlap between highlighted genes and genesets will not be guaranteed even if DNA methylation changes identified are aetiologically relevant. When DNA methylation mediates the effect of genetic variants distal to their genomic position on complex traits the genes identified by GWAS and EWAS will also differ, but the genesets would likely overlap. A lack of overlap could also reflect the fact associations in EWAS may be driven by confounding and reverse causation. Despite the caveats mentioned, it is plausible that in the absence of confounding and reverse causation genes and genesets identified in EWAS would overlap more than expected by chance with those identified by GWAS. The extent to which this expectation holds is explored in more detail in the **Discussion**.

In this paper we determine the overlap between genes and genesets identified by GWAS and EWAS of 14 complex traits and we use these results to infer genetic and epigenetic architectures that are likely to be consistent with our results. In this paper we do not attempt to infer the causal structure of DNA methylation-trait associations directly as has been attempted through methods such as Mendelian randomization, but is often difficult due to being unable to prove the vertical pleiotropy assumption (16,22). Rather, we adopt a different approach by simulating a range of causal systems to explore potential interpretations of the patterns of overlap between genomic features reported by EWAS and GWAS. For the purposes of this study we denote the terms “causal gene” as a gene that affects the trait of interest and an “associated gene” as a gene that correlates with the trait of interest, but might not affect it (**Figure 1**).

**Figure 1:**
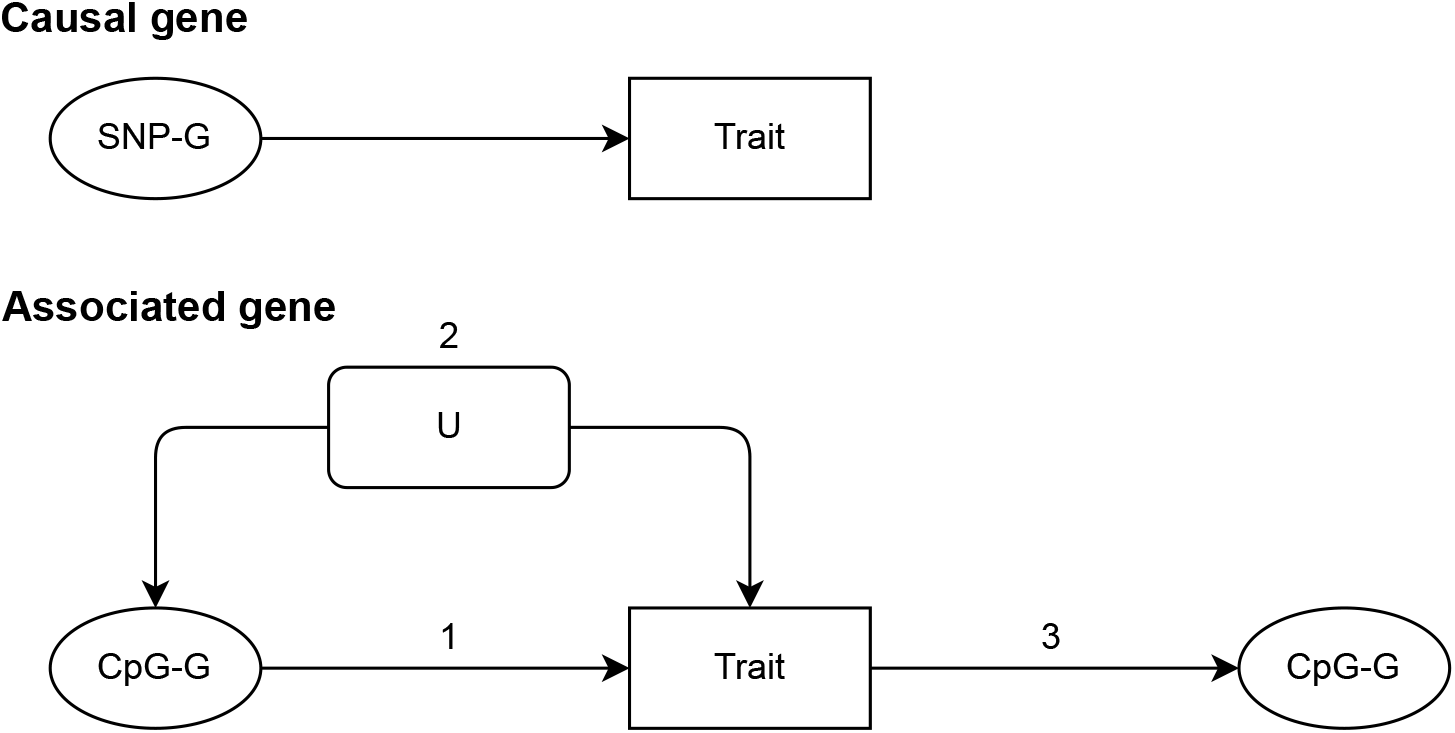
A diagram of causal and associated genes. A causal gene is one where the product of that gene affects the trait of interest and in this study we are assuming that SNPs identified in relation to a trait will affect these genes or tag SNPs that do (SNP-G). An associated gene is one where the product of that gene correlates with the trait of interest, but may not affect it. In this study we are assuming that CpG sites identified in relation to a trait will map to these genes (CpG-G). The diagram shows how a gene product may be correlated with a trait: 1. by affecting the trait, 2. by sharing a common cause with the trait (confounding), 3. by being affected by the trait (reverse causation). A geneset may be made of causal and associated gene products. U = confounder.

## Results

### Study data

We selected traits for which EWAS had been conducted with over 4500 samples, had more than 10 associations at P<1×10^−7^ and for which corresponding well-powered GWAS summary data were available. Suitable traits were identified using The EWAS Catalog (23) on 2021-03-01. Traits and study data information is in **Table 1**.

**Table 1:** Study data

| trait | EWAS author | EWAS PMID | EWAS n | GWAS author | GWAS PMID | GWAS n |
| --- | --- | --- | --- | --- | --- | --- |
| body mass index | Wahl S | 28002404 | 10,238 | Yengo L | 30124842 | 681,275 |
| current versus never smoking | Joehanes R | 27651444 | 9,389 | Liu M | 30643251 | 632,802 |
| former versus never smoking | Joehanes R | 27651444 | 13,474 | Elsworth B | NA | 424,960 |
| alcohol consumption per day | Liu C | 27843151 | 9,643 | Liu M | 30643251 | 335,394 |
| c-reactive protein | Ligthart S | 27955697 | 8,863 | Ligthart S | 30388399 | 204,402 |
| educational attainment | Karlsson Linner R | 29086770 | 10,767 | Lee JJ | 30038396 | 766,345 |
| chronic kidney disease | Chu A. | 29097680 | 4,859 | Pattaro C | 26831199 | 117,165 |
| fasting glucose | Liu J | 31197173 | 4,808 | Manning AK | 22581228 | 58,074 |
| fasting insulin | Liu J | 31197173 | 4,808 | Manning AK | 22581228 | 51,750 |
| systolic blood pressure | Richard MA | 29198723 | 17,010 | Evangelou E | 30224653 | 757,601 |
| diastolic blood pressure | Richard MA | 29198723 | 17,010 | Evangelou E | 30224653 | 757,601 |
| birthweight | Kupers L | 31015461 | 8,825 | Horikoshi M | 27680694 | 143,677 |
| cognitive abilities: digit test | Marioni R | 29311653 | 4,794 | Lee JJ | 30038396 | 257,841 |
| fev1 | Imboden M | 31073081 | 5,370 | Elsworth B | NA | 421,986 |

PMID = PubMed ID, author = first author on the publication.
Where GWAS PMID = NA, the GWAS were conducted as part of a UK Biobank GWAS pipeline within the University of Bristol's Integrative Epidemiology Unit and can be found on the OpenGWAS Project website (see Methods for more).

### Genomic position overlap

We divided the genome into 5591 non-overlapping 500kb regions and mapped EWAS and GWAS signals for each trait to these regions (See **Methods** for more details). For each trait, the number of regions that were identified by one study type and not the other was higher than the number of overlapping regions (**Figure 2**). Further, the magnitude of the greatest GWAS effect estimate in each region had little ability to predict whether or not a DNA methylation site was likely to be identified in the same region (AUC range = 0.43-0.61, **Supplementary figure 1**).

**Figure 2:**
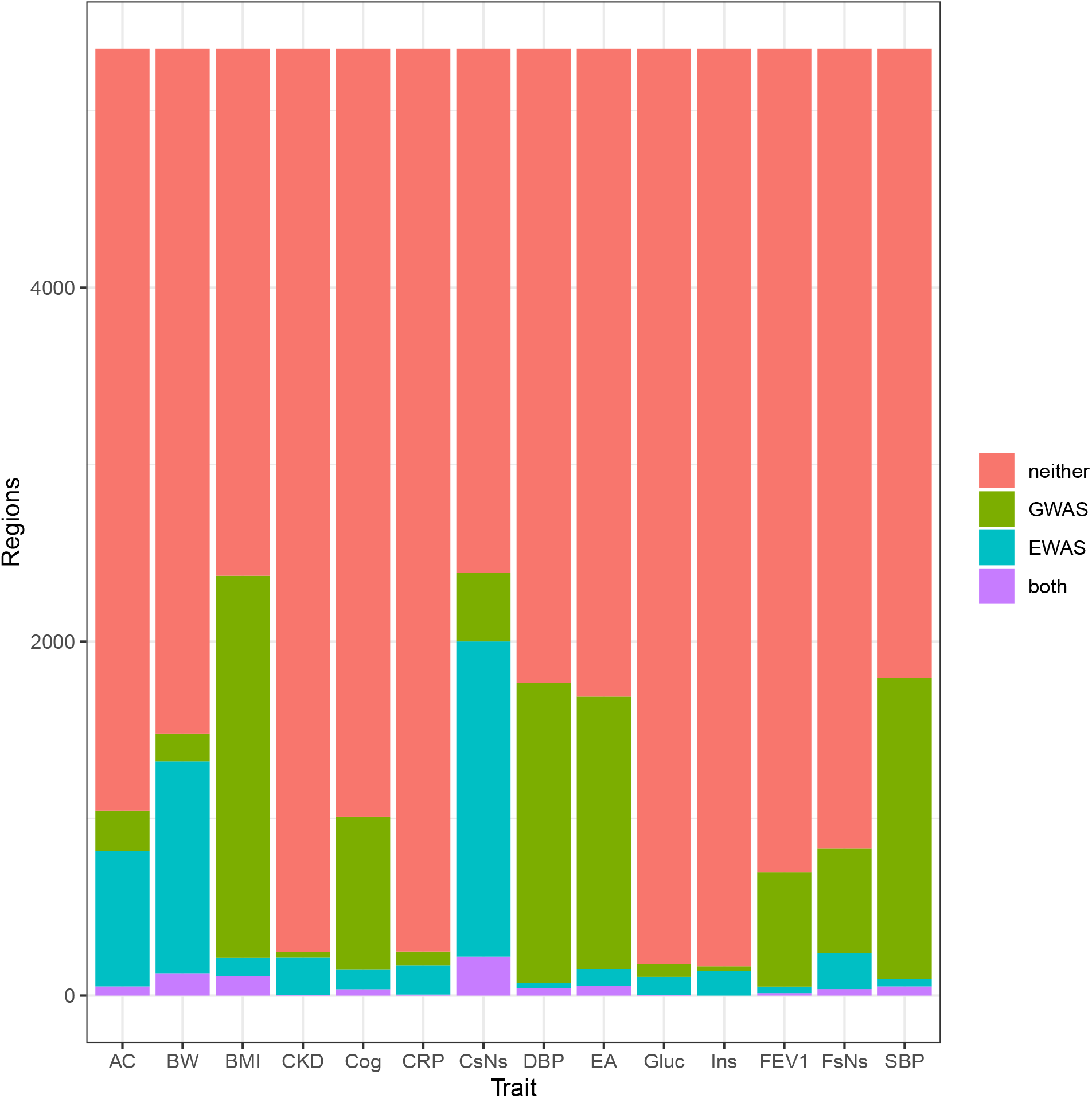
Overlap between genomic positions identified by corresponding EWAS and GWAS. The genome was divided into 500Kb regions. Those where no probes on the HM450 array measured DNA methylation were excluded from the analysis. Regions were counted as being identified by a GWAS if one or more SNPs in that region associated with the trait and as being identified by an EWAS if one or more CpGs in that region associated with the trait. Neither = no EWAS or GWAS sites identified in the region, GWAS = GWAS sites only were identified, EWAS = EWAS sites only were identified, Both = Both EWAS and GWAS sites were identified, AC = alcohol consumption per day, BW = birthweight, BMI = body mass index, CKD = chronic kidney disease, Cog = cognitive ability (digit test), CRP = c-reactive protein, CsNs = current smokers vs never smokers, DBP = diastolic blood pressure, EA = educational attainment, Gluc = fasting glucose, Ins = fasting insulin, FEV1 = forced expiratory volume in one second, FsNs = former smokers vs never smokers, SBP = systolic blood pressure.

### Assessing power to detect shared annotations between GWAS and EWAS

Genomic function and trait biology are not divided into discrete 500kb genomic chunks, thus GWAS and EWAS could still be identifying similar facets of trait biology without identifying the same genomic regions.

We sought to assess whether the genes and genesets identified overlapped more than expected by chance, thus genomic positions were mapped to genes and genes to genesets (details in **Methods**). Overlap between genes identified was assessed using Fisher’s exact test. Two methods for testing overlap between genesets were considered, one simply mapped genes to genesets and used Fisher’s exact test in the same way as assessing overlap between identified genes. The other generated ‘enrichment scores’ for each geneset and assessed correlation between the geneset enrichment scores across studies.

In the case that genetic influences on disease are mediated via cis-acting DNAm levels, or other proximal regulatory factors that in turn correlate with proximal DNAm levels, if all DMPs that associated with a trait were on the pathway from SNP to disease, then EWAS and GWAS would be identifying genes from the exact same geneset (i.e. genes that caused changes in the trait). The more DMPs that are identified because of confounding effects or reverse causation, the smaller the chance of overlap, assuming that causal and responsive genesets are independent (more on this in the **Discussion**). In a scenario where no DMPs are causing phenotypic changes then any overlap in genes and genesets found would be entirely attributable to chance. We ran simulations to assess which scenarios the enrichment and annotation methods had power to detect whether there was more overlap than expected by chance. Power was also assessed across different annotation methods. A schematic of how the simulations were set up can be found in **Supplementary figure 2**.

Under each of a range of genetic and epigenetic architectures and study sizes, the ability to infer whether EWAS were identifying, at least in part, the same set of genes as GWAS (‘causal genes’) compared to a random set of genes (‘associated genes’) was tested. Performance improved as the study sample sizes and the proportion of DMPs that were causal increased (**Figure 3, Supplementary figure 3**). Performance of the overlap tests tended to increase as the number of identified genes increased, but this parameter was largely inconsequential when the proportion of DMPs that were causal was low (**Supplementary figure 3**). Performance was similar across annotation methods and between methods attempting to assess geneset overlap, with assessment of gene overlap performing better (**Figure 3**). Overall there was more power to detect overlap in genes than overlap in genesets. Between the geneset methods, there was more power to detect correlation between enrichment scores than direct overlap in genesets. Therefore, gene overlap and correlation between geneset enrichment scores were taken forward for the empirical analyses. Assigning genes to GO terms (24,25) to use as genesets had more power, than assigning genes to genesets derived from the KEGG (26–28), Reactome (29) or protein-protein interaction database from EpiGraphDB (30), when assessing overlap between these genesets (**Figure 3**). Therefore, GO terms were taken forward and used as the geneset annotations in the empirical analyses.

**Figure 3:**
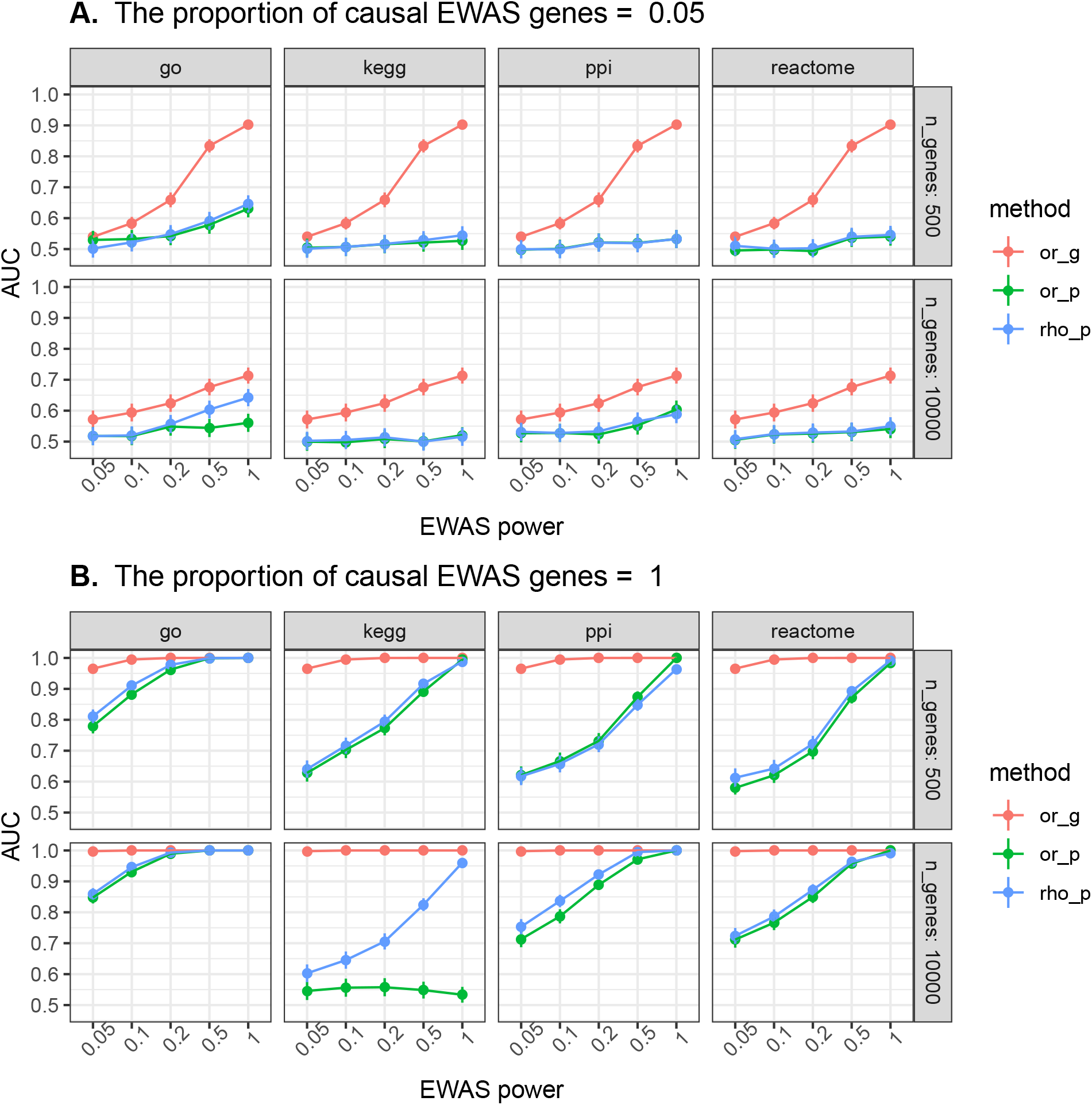
Power to detect overlap between genes and genesets identified by corresponding EWAS and GWAS. Simulations were set up as illustrated in **Supplementary figure 2**. EWAS power is equivalent to the proportion of “n_genes” EWAS is detecting. In the scenario where n_genes = 500, EWAS power = 1, and the proportion of causal EWAS genes = 0.05, the EWAS is detecting 500 genes, 25 of which are causal. The area under receiver operator curves (AUC) was used to estimate the ability to distinguish between results generated when EWAS and GWAS were sampling, in part, from the same set of causal genes and results generated when EWAS was sampling random genes from the genome. The header of each set indicates the proportion of genes identified by the simulated EWAS that were set to be causal. or_g = assessing overlap of genes, or_p = assessing overlap of genesets, rho_p = assessing correlation between geneset enrichment scores. go = gene ontology, ppi = protein-protein interaction database from EpiGraphDB. This is a summary of the results, full results can be found in **Supplementary figure 3**.

### Gene and geneset overlap between GWAS and EWAS

For the 14 traits used, the number of genes identified by EWAS and GWAS that overlapped was low and for four traits no genes identified by the studies overlapped (**Table 2**). The number of genesets that overlapped was higher, peaking at 1,243 for GWAS and EWAS of body mass index (**Table 3**).

**Table 2:** Overlap of genes identified by EWAS and GWAS

| trait | n EWAS genes | n GWAS genes | gene overlap | obs OR | exp overlap | exp OR | p diff |
| --- | --- | --- | --- | --- | --- | --- | --- |
| body mass index | 232 | 3,221 | 23 | 2.0 | 39 | 3.04 | 5.2e-02 |
| current versus never smoking | 1,933 | 312 | 27 | 2.9 | 39 | 3.42 | 3.2e-01 |
| former versus never smoking | 282 | 320 | 9 | 6.4 | 7 | 3.43 | 2.2e-02 |
| alcohol consumption per day | 361 | 190 | 3 | 2.6 | 3 | 2.44 | 9.1e-01 |
| c-reactive protein | 189 | 302 | 3 | 3.2 | 1 | 0.91 | 1.2e-01 |
| educational attainment | 25 | 1,594 | 1 | 1.5 | 3 | 3.00 | 4.3e-01 |
| chronic kidney disease | 48 | 11 | 0 | 0.0 | 0 | 0.00 | 1.0e+00 |
| fasting glucose | 15 | 50 | 0 | 0.0 | 0 | 0.00 | 1.0e+00 |
| fasting insulin | 36 | 5 | 0 | 0.0 | 0 | 0.00 | 1.0e+00 |
| systolic blood pressure | 99 | 3,210 | 18 | 4.0 | 12 | 2.13 | 9.4e-03 |
| diastolic blood pressure | 56 | 3,586 | 15 | 5.8 | 6 | 1.87 | 5.2e-06 |
| birthweight | 894 | 161 | 6 | 2.6 | 7 | 2.64 | 9.1e-01 |
| cognitive abilities: digit test | 26 | 875 | 0 | 0.0 | 1 | 2.44 | 4.2e-01 |
| fev1 | 51 | 1,657 | 4 | 3.0 | 3 | 1.92 | 3.9e-01 |

exp = expected, obs = observed, fev1 = forced expiratory volume in 1 second
odds ratios (ORs) can be interpreted as the odds of an gene being identified by EWAS and a GWAS over the odds of a gene being identified by an EWAS but not by a GWAS.
exp OR = the mean OR after repeating the analysis 1000 times, randomly sampling EWAS genes equal to the number identified in the empirical analysis.
p diff = P value from assessing the difference between the observed and expected OR.

**Table 3:** Correlation of geneset enrichment scores between EWAS and GWAS

| trait | n ewas genes | n gwas genes | geneset overlap | obs cor | exp cor | p diff |
| --- | --- | --- | --- | --- | --- | --- |
| body mass index | 232 | 3,221 | 1,243 | 0.187 | 0.20 | 0.2287 |
| current versus never smoking | 1,933 | 312 | 1,053 | 0.200 | 0.22 | 0.0343 |
| former versus never smoking | 282 | 320 | 661 | 0.298 | 0.30 | 0.9936 |
| alcohol consumption per day | 361 | 190 | 562 | 0.259 | 0.27 | 0.5753 |
| c-reactive protein | 189 | 302 | 600 | 0.265 | 0.26 | 0.7983 |
| educational attainment | 25 | 1,594 | 215 | 0.105 | 0.13 | 0.1729 |
| chronic kidney disease | 48 | 11 | 31 | 0.131 | 0.14 | 0.6852 |
| fasting glucose | 15 | 50 | 50 | 0.153 | 0.16 | 0.7723 |
| fasting insulin | 36 | 5 | 19 | 0.093 | 0.12 | 0.4400 |
| systolic blood pressure | 99 | 3,210 | 631 | 0.145 | 0.14 | 0.7679 |
| diastolic blood pressure | 56 | 3,586 | 594 | 0.160 | 0.11 | 0.0013 |
| birthweight | 894 | 161 | 683 | 0.224 | 0.24 | 0.2476 |
| cognitive abilities: digit test | 26 | 875 | 198 | 0.164 | 0.14 | 0.2064 |
| fev1 | 51 | 1,657 | 403 | 0.158 | 0.14 | 0.1948 |

For each geneset, odds of study genes being in the geneset divided by the odds of the study genes not being in the geneset were assessed and correlation between these odds ratios are given here.
exp cor = the mean correlation between odds ratios after repeating the analysis 1000 times, randomly sampling EWAS genes equal to the number identified in the empirical analysis.
geneset overlap indicates the number of gene ontology terms that map to both genes identified by the EWAS and GWAS.
p diff = P value from assessing the difference between the observed and expected correlations.

The number of overlapping genes identified was no more than expected by chance for 12 of 14 traits, but for systolic blood pressure and diastolic blood pressure, the overlap between observed genes was greater than expected (6 more genes and 9 more genes overlapped than expected by chance respectively) (**Table 2**). There was also evidence that correlation between enrichment scores of the GO terms identified by GWAS and EWAS of diastolic blood pressure was greater than expected by chance (P = 0.0013), but there was little evidence for this across the other traits, including systolic blood pressure (**Table 3**).

There were 56 GO term genesets that were commonly enriched (FDR < 0.1) for both the EWAS and GWAS traits. Of these zero were specific (contained under 100 genes). There were 261 specific genesets (geneset size < 100 genes) that did not overlap between studies of corresponding traits, for example, the genes identified by the GWAS of alcohol consumption were enriched for the “ethanol catabolism” pathway (geneset size = 12 genes), however none of the genes identified by the EWAS were present in this pathway.

### Understanding architecture from geneset overlap

Given observations of numbers of genesets discovered and geneset overlap for a particular trait, we next asked if we can impose bounds on the likely genetic and epigenetic architectures of that trait. To do this we assumed discovered genes and genesets represented some fraction of total genes and genesets, and then re-sampled EWAS and GWAS results to ascertain architectures that generated overlap scores matching the empirical results.

For the simulations, three sets of genes were linked to each trait: genes identified by the GWAS (known GWAS genes), genes identified by the EWAS (known EWAS genes) and a random set of genes sampled from the total set of Ensembl gene IDs (excluding the genes identified by the EWAS and GWAS).

Having generated the GWAS and EWAS genes, enrichment of GO terms was performed and the correlation between enrichment scores across all the terms was estimated. A schematic of the methods for these simulations can be found in **Supplementary figure 4** and it is described in full in the **Methods**.

From the simulations, we determined, that for most traits the overlap between causal and associated genes is unlikely to be high if the total number of genes still to discover for these traits is low. However, for some traits such as systolic blood pressure, diastolic blood pressure and former vs. never smoking the simulations suggest the overlap of causal and associated genes is high. It should be noted though, that the correlation of enrichment scores across different scenarios varied little for each trait, making inference difficult. The results from the analysis of former vs. never smoking and c-reactive protein (representing simulations for the other traits) are shown in **Figure 4. Supplementary Figure 5** shows the same results for the other traits. Each simulation was repeated 1000 times. Evidence for a difference in the empirically determined correlation of geneset enrichment scores and the mean correlation of geneset enrichment scores across simulations was assessed using a z-test for difference. There was some evidence against 115 simulation scenarios (FDR < 0.05). Across the traits, the scenarios that were least likely tended to be when the number of genes yet to discover was low, and the overlap between causal and associated genes was high, except for former vs. never smoking, highlighting architecture differences between traits.

**Figure 4:**
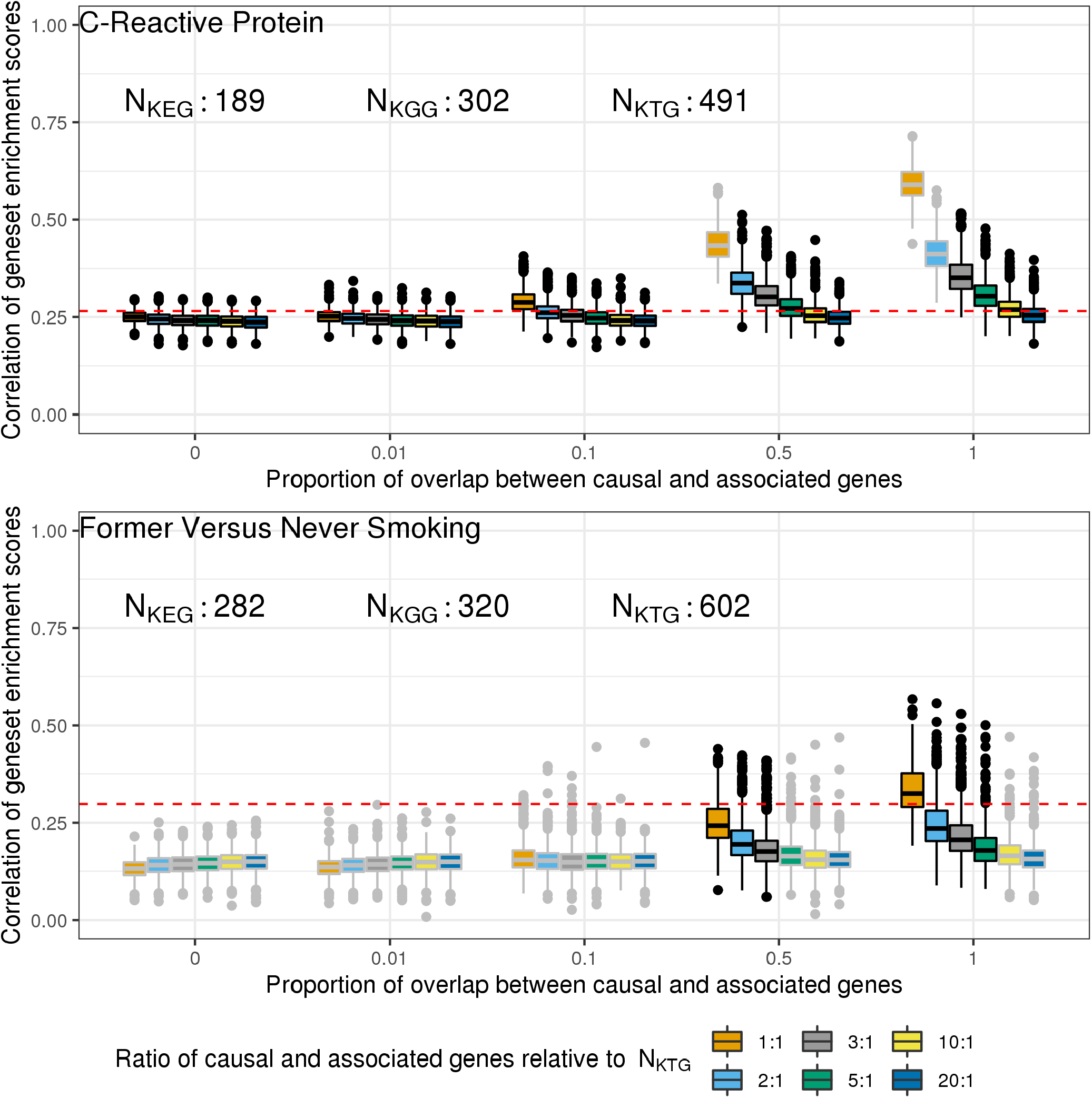
Simulations to understand the likely number of genes still to identify in EWAS and GWAS of C-reactive protein and smoking (former vs. never smokers) under different trait architectures. Simulations were set up as illustrated in **Supplementary figure 4**. Correlation of geneset enrichment scores from empirical data (**Table 3**), is shown as a red dashed line. Box plots show the range of enrichment score correlations from 1000 simulations using the parameters indicated. The number of causal and associated genes, as well as the overlap between these genes were varied. N_KEG_ = number of known EWAS genes, N_KGG_ = number of known GWAS genes, N_KTG_ = number of known total genes (N_KEG_ + N_KGG_). By way of an example, when N_KTG_ = 557 and the ratio of causal and associated genes relative to N_KTG_ is 1:1, the number of causal genes in the simulations will be 557 and the number of associated genes in the simulations will be 557. Scenarios which lie close to the empirical result (red dashed line) are more likely to reflect the true underlying number of genes related to a trait and the true overlap between the causal and associated genes. Therefore, for both current vs. never smoking and for body mass index, one could say that the overlap of causal and associated genes is likely to be below 0.5, unless there are many more genes to identify in EWAS and GWAS.

### Overlap of non-corresponding EWAS and GWAS

We next hypothesised that EWAS of one trait might actually relate more closely to GWAS of another trait. For example, if DNA methylation changes related to BMI were mediating the effect of BMI on changes in metabolites, then one could expect to see greater correspondence in identified genes and genesets with GWAS of those metabolites.

To test if this was the case for any of the EWAS in this study we extracted 1886 GWAS with at least one SNP associated at P < 5×10^−8^ with a sample size above 5000 from the IEU OpenGWAS Project (31,32). For each GWAS and the 14 EWAS, enrichment scores were calculated using geneset enrichment analysis and correlation between the enrichment scores was calculated, as in previous analyses.

Across all pairwise comparisons, correlations between enrichment scores ranged from -0.014 to 1 and had a mean of 0.12 (**Supplementary figure 6**). The mean correlation between GWAS traits and EWAS traits was 0.26, which was higher than the correlations between just GWAS traits (0.12). Amongst just GWAS results, there was evidence that 14652 pairwise enrichment score correlations were greater than the the mean (FDR < 0.05). However, there was little evidence that any pairwise correlations between enrichment scores derived from EWAS and GWAS were greater than the mean correlation (FDR > 0.05).

This suggests that the signal from EWAS is not capturing aspects of any specific factor that impacts the aetiology of any of the 14 traits of interest.

## Discussion

Several EWAS papers have compared their findings with those of the corresponding trait GWAS (6,33–36), but it’s unknown if any overlap that might occur should be attributed to shared underlying architectures or if it occurs by chance. In this study, the genes and genesets identified by 12 of 14 large EWAS (N > 4500) were not identified in their corresponding GWAS any more than expected by chance, with the other two being highly similar phenotypes, systolic and diastolic blood pressure. Simulations suggested these EWAS could still be identifying aspects of trait aetiology, but it’s likely most DMPs identified are due to confounding or reverse causation. Further simulations suggested that the overlap between genes that impact phenotypic variation and those that might be identified through confounded analyses or because of reverse causation is likely to be low amongst EWAS of most traits. However, if the number of genes still to identify in EWAS and GWAS is high, it is possible that the overlap between “causal” and “associated” genes could be high also.

### Overlap expected

GWAS identifies the effects of genetic variation on complex traits. These effects are less likely to be confounded than associations estimated between observational phenotypes (37,38). Thus, one would expect overlap between genes and gemesets identified by EWAS and GWAS of the same trait if the DMPs identified are also of aetiological relevance. Assuming mapping of DMPs and SNPs to genes is correct, the genes identifed by EWAS may cause variation in complex traits without overlapping with genes identified in GWAS. Under the scenario where the effect of a SNP on a complex trait is mediated by a distal DNA methylation site (or the gene that site is tagging), GWAS and EWAS may identify genes that do not overlap, but that are along the same causal pathway from genome to complex trait. One plausible mechanism for which trans-mQTLs may act is via transcription factors. A trans-mQTL may influence the transcription of a nearby transcription factor then the transcription factor could cause a change in DNA methylation at distal sites. One study has provided evidence this may occur frequently (39). In this scenario the genes proximal to the identified SNP and DMP would lie on the same causal pathway and so would likely be part of the same genesets. This is likely not the only plausible mechanism of trans-mQTL function though. DNA methylation may also mediate non-genetic effects, allowing for causal DMPs to be identified at genes not near pertinent genetic variation. However, there is strong evidence that the majority of DNA methylation sites have a heritable component. As such any effect of DNA methylation on a trait could be influenced by genetic variation. If DNA methylation is influenced by proximal genetic variants then the discovery of the same gene(s) will be a function of GWAS, EWAS power and the heritability of the DNA methylation sites. If only distal genetic variants influence the DNA methylation site, then the overlap of genesets is a function of regulatory mechanisms and power. These two issues, and likely others, may introduce some noise into the results. However, there is overwhelming evidence that confounding and reverse causation are pervasive across observational epidemiology (40–43) as well as within EWAS (3,44). This suggests that the evidence that EWAS and GWAS are not identifying any more overlapping genes and genesets than expected by chance, is likely due to identified DMPs being mostly being the result of reverse causation or confounding.

As the simulations showed, even if an EWAS identifies DMPs that cause a change in the trait, if the majority of DMPs identified are due to confounding or reverse causation then the overlap will be indistinguishable to the overlap expected by chance. Thus, our empirical results do not preclude the possibility that some DMPs identified by the EWAS are involved in trait aetiology.

For body mass index, work has already suggested the trait causes changes in DNA methylation rather than *vice versa* (17), supporting our findings here. Further, one study suggested DNA methylation changes capture different components of body mass index variance than genetic variation (45) and another estimated the percentage of trait variance captured by DNA methylation was 76% when accounting for genotype (46).

Simulations involving empirical data from former vs. never smoking EWAS and GWAS suggested that overlap between causal and associated genes was high. This is surprising for two reasons. Firstly, it differs from the current vs. never smoking results, suggesting distinct genetic or epigenetic architectures of those traits. Secondly, there is evidence that for DNA methylation changes identified in relation to smoking, smoking is likely causing DNA methylation variation and not *vice versa* (47). Although the statistical tests suggested that it was unlikely that the proportion of causal and associated genes is 0.1 or lower, it should be noted that the absolute difference between simulated results and empirical results were not great.

It’s also potentially unexpected that genes identified by EWAS diastolic and systolic blood pressure overlapped more with genes identified by their respective GWAS than expected by chance. In the study that conducted the EWAS of both blood pressure phenotypes, the authors conducted bi-directional Mendelian randomization analyses to improve causal inference of DNA methylation changes identified in the EWAS (48). They presented evidence that variation in blood pressure causes changes in DNA methylation at 4 sites, and evidence for the reverse at one site. This suggests that the majority of DNA methylation sites identified in their EWAS are due to reverse causation. However, not all CpG sites could be instrumented and not all sites identified in the EWAS were taken forward for the Mendelian randomization analyses. Further, for the sites taken forward, none had a large number of genetic instruments, making evaluation of pleiotropy difficult. Therefore, it is plausible that some of the sites identified in their EWAS are tagging causal genes and this could not be tested by the authors. In addition to this, a GWAS of blood pressure found that identified SNPs were enriched for association with DNA methylation at CpG sites within 1Mb (49). Similar, to our study, this work also suggests that DNA methylation variation is tagging causal blood pressure loci, without formally testing whether DNA methylation changes effect the trait directly.

If EWAS is discovering some DMPs that influence genes that cause changes in the trait of interest, study power is the limiting factor for detecting overlap. For traits with a weak polygenic architecture (few genes explain most of the heritability), such as gene expression (50,51), discovering almost total overlap would be inevitable even with modest sample sizes.

### Little overlap with any GWAS

Little correlation was found between geneset enrichment scores for EWAS and GWAS of different traits. In a scenario where DNA methylation was capturing a specific facet of a trait one might expect correlation between EWAS of the original trait and GWAS of that facet. For example, if changes in DNA methylation associated with smoking were mostly responsible the effect of smoking on lung cancer then one would expect to observe an overlap between the genes and pathways identified by an EWAS of smoking and GWAS of lung cancer. This specific example has been examined before, with studies suggesting either methylation at two sites (of over 1000 smoking-related sites) mediate over 30% of the effect of smoking on lung cancer (52) or that there is little evidence for a causal effect of DNA methylation on lung cancer (11). The results of either study suggest most sites will not mediate the effect of smoking on lung cancer and thus there would be little overlap between genes and pathways of an EWAS of smoking and a GWAS of lung cancer. This is corroborated by the results of our study: there was little evidence that correlation of pathway enrichment scores between the two, 0.15, was greater than the mean correlation enrichment score across all GWAS-GWAS and GWAS-EWAS correlations, 0.12 (FDR > 0.05).

It is important to note that overlap between EWAS and GWAS genesets may be missed even if this mediation model is true for various traits. As shown in the simulations, detecting this overlap depends on individual study power as well as the underlying genetic architecture of the trait. There are further things that may limit detection of geneset overlap that are discussed in the limitations below.

### Information gained from EWAS

The fact that genes and pathways identified from most GWAS and EWAS of the same traits are seemingly very separate suggests we are gaining new information from EWAS, even if interpreting the new information may be difficult. Key to interpreting the EWAS results would be to try and disentangle whether the EWAS results are likely due to confounding. Interpreting EWAS can also be difficult due to cell type heterogeneity and the complexity of mechanisms which mediate DNA methylation changes (3,8,53–55). Due to these difficulties, it should not be concluded that EWAS definitely help increase our biological understanding of complex traits. Rather, DNA methylation is capturing different biological information. Regardless of biological insight gained, translational impact may still be gleaned from DNA methylation studies; DNA methylation may aid diagnoses by acting as a reliable biomarker or could help predict various health outcomes (2).

There are also benefits to understanding the biological consequences of a trait, something that EWAS might help identify and GWAS will not (at least not directly). This does depend on further research to understand how changes in DNA methylation downstream to complex trait variation is relevant to human health. Further, establishing where exactly DNA methylation may lie on the causal pathway may be difficult and work is ongoing to discover this for various traits (22). Use of causal inference methods such as Mendelian randomization (37,38,56) have been applied, but this still comes with various caveats (22,56). Some studies also try and confirm effects experimentally (1,11) and use previous biological knowledge of the trait to try and understand EWAS results.

However, some of the biological knowledge of complex traits relating to genes and pathways comes from GWAS of those traits. This study suggests that EWAS is unlikely to identify many genes proximal to genetic variation pertinent to the trait of interest and further the genesets are unlikely to overlap with those identified in GWAS. Therefore, comparison of EWAS results to those of a corresponding GWAS is unlikely to yield much insight. This may make inference from EWAS difficult, yet it seems likely the interpretability of DNA methylation studies will continue to improve over the coming years, as understanding the underlying epigenetic architecture of complex traits could still provide translational benefits (2).

### Limitations

As discussed, detecting gene or pathway overlap depends on the genetic and DNA methylation architecture of the trait. Here only 14 traits, two of which are smoking behaviour traits and two of which are blood pressure traits, have been studied. Further, these traits are mostly exposures that precede disease. This means the results cannot be generalised to all or even the majority of complex traits or diseases. These analyses could be repeated by setting a less restrictive sample size limit, but it was felt that would make the results less reliable and impossible in many circumstances where too few DNA methylation sites had been discovered by EWAS. As sample sizes increase and technologies measuring more DNA methylation sites become more common, it would be interesting to repeat the analysis.

Often in GWAS and EWAS, prioritisation of SNPs and DMPs identified occurs before functional mapping. Prioritisation for both studies may be informed by prior knowledge of the trait, prior understanding of molecular biology, predicted consequences of observed variation (for example Ensembl’s Variant Effect Predictor (57)), replication of findings or a number of other methods. In this study, we did not perform any prioritisation (besides the conventional P value threshold cutoffs) and thus may have increased the amount of “noise” in the signal taken forward for functional annotation. Unfortunately, this extra prioritisation of sites is not tractable when comparing many different association studies and may reduce power to detect any overlap between genes and pathways. However, added noise is unlikely to prevent detection of true overlap if that true overlap is substantial, as shown by our simulations (**3**).

The nearest gene, by chromosomal position, to a DMP or SNP is not necessarily the gene of interest. SNPs may have effects on genes distal to their position (58) and the correlation between genetic variants inflates associations of variants proximal to the true causal variant, which may map to unrelated genes. Further, the correlation structure in DNA methylation data may induce associations between complex traits at a site far from where variation in DNA methylation causes complex trait changes (58). Therefore, the mapping of DMPs and SNPs to genes in this study could likely be improved. The “correct” method for this mapping has not yet been established though and even though some tools are available (such as eQTL studies), there are caveats to them too (59,60).

Our understanding of molecular pathways is not complete and thus attributing genes to certain pathways or functionalities may be erroneous. However, the results remained consistent across four different methods that annotate genes to pathway, suggesting differences in mapping genes to genesets should not impact our conclusions.

Biological information gained from EWAS and GWAS may be defined in various ways and depending on the interpretation of this, one could alter methods used to extract biological information. However, first exploring the genomic regions identified and then mapping these to potentially relevant biological pathways is common amongst GWAS and EWAS (4–7,61–63) and provided a simple way to compare the information from the two study types.

A simple explanation of EWAS genes not overlapping with GWAS genes is that they are not causal. However, it is possible that GWAS genes are limited to a small subset of causal genes in which functional variation is evolutionarily permissible. Because epigenetic variation can be modified by the environment it’s possible that causal genes are that are unavailable to GWAS through lack of functional variation are identified through EWAS. Such a scenario would reduce power to detect overlap using the simulation approaches that we employed here.

## Conclusion

Overall, this study provides evidence that, for 13 of 14 complex traits, there is little overlap between genes and genesets identified by EWAS and GWAS of the same trait. Given the differences in properties between DNA methylation and genetic variants the results presented in this study may apply to other traits, but this is still to be confirmed. Where lack of overlap between genes and genesets is found, it suggests EWAS may be providing new biological information, however, the interpretability of EWAS is still in question and with current methods it is hard to determine if EWAS results are attributable to confounding or reverse causation. Regardless, as datasets grow and causal inference methods improve we are likely to be better able to interpret the role of DNA methylation in complex traits.

## Methods

### Samples

EWAS summary data and GWAS summary data were extracted from The EWAS Catalog (23) and the IEU OpenGWAS Project (31,32) respectively. For traits that had multiple EWAS with a sample size of greater than 4500, the EWAS with the largest sample size was used, the same was applied to the GWAS. The sample size, first authors and PubMed IDs can be found in **Table 1**.

### Overlapping genomic regions

Each chromosome was divided into 500Kb blocks, each block that did not contain a DNA methylation site measured by the Illumina Infinium HumanMethylation450 BeadChip was removed. For each trait, the genome blocks that had one or more EWAS sites and one or more GWAS sites that reached the set p-value threshold were tallied. The p-value threshold was set at a lenient P<1×10^−5^ or if it was lower, the maximum reported p-value in the EWAS of that trait.

### Mapping sites to genes and genesets

The R package biomaRt (64) was used to extract Ensembl gene ids along with chromosome positions of all genes. The package was also used to extract gene ontology (GO) terms (24,25) and map these to the Ensembl gene ids. The R package limma (65) was used to extract KEGG terms (26–28) and these were mapped to Ensembl gene ids.

Protein-protein interaction data, which includes data from StringDB (66) and IntAct (67), and terms from the Reactome database (29) were extracted from EpiGraphDB (30).

CpG sites associated with traits at P < 1×10^−7^ and SNPs associated with traits at P < 5×10^−8^ were taken forward to be mapped to genes. The correlation structure present in genetic and DNA methylation data makes it difficult to ascertain the precise site driving any signal observed. Thus, no filtering based on correlation between variants or CpG sites was performed.

For each CpG site identified by EWAS and used in the analyses, it was mapped to the nearest gene (Ensembl gene ID) by chromosome position. If a CpG site lay within the bounds of multiple genes then the site was mapped to all of those genes. Therefore, one CpG site could map to multiple genes and one gene could map to multiple CpG sites. The same gene mapping approach was used for variants identified in GWAS. The positions of CpG sites were extracted using the R package meffil (68).

### Methods for assessing overlap

To test the overlap between genes identified we generated ORs as so

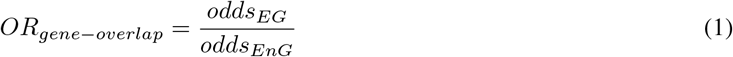

where *odds*_*EG*_ is the odds of a gene being identified in EWAS and GWAS and *odds*_*EnG*_ is the odds of a gene being identified in EWAS, but not in GWAS.

Genes may map to genesets by chance. Often in EWAS and GWAS, enrichment for any genesets are tested by assessing whether the genes identified are more common in any geneset than expected by chance. We tested mapping genes to genesets and directly assessing overlap like in Equation (1) against correlation between enrichment scores for each geneset. Enrichment scores are also odds ratios generated in a similar way to those in Equation (1):

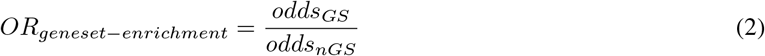

where *odds*_*GS*_ is the odds of a gene being annotated to the geneset and *odds*_*nGS*_ is the odds of a gene not being annotated to the geneset.

For many genesets, genes annotated to that geneset would not be identified in an EWAS or GWAS, causing the enrichment score for many genesets to be zero. Further, some genesets would have very large enrichment scores. This made the relationship between enrichment scores generated for the EWAS results and the enrichment scores generated for the GWAS non-normal. Thus, to examine the relationship between the two, we generated Spearman’s rank correlation coefficients for the logarithm of enrichment scores.

### Testing power to detect overlap

Simulations were setup as seen in **Supplementary figure 2**. The simulations iterated over each set of parameters 1000 times. For each iteration, two sets of genes, GWAS genes and EWAS genes, were sampled from the total set of genes. Each iteration assessed gene overlap and geneset overlap between these gene sets using Equation (1). Equation (2) was used to generate enrichment scores for each gene set and then correlation between the enrichment scores was assessed.

GWAS genes were only sampled from a set of “causal” genes and a proportion of EWAS genes were sampled from the set of causal genes and the rest from the set of “associated” genes. Receiver operator characteristic (ROC) curves were generated to assess whether it was possible to predict the gene overlap, geneset overlap and enrichment score correlations for scenarios where the proportion of causal EWAS genes was greater than zero from the scenario where the proportion of causal EWAS genes was zero. The area under these ROC curves were then calculated in each case.

These simulations were repeated for each geneset and protein-protein interaction database. The protein-protein interaction and Reactome databases all map only to protein coding genes, whereas the GO and KEGG databases map to all Ensembl gene IDs. To compare predictive ability across annotation methods, we excluded Ensembl gene IDs that were not protein coding genes. We also compared the performance of models when mapping to all Ensembl gene IDs and protein coding genes only for GO and KEGG databases. (**Supplementary figure 7**).

From these simulations, the best method to assess geneset overlap, and the best geneset annotation method to assess that overlap and the scenarios (i.e. study power required, proportion of DMPs that need to be causal) in which we expect to be able to detect overlap could be deduced.

### Empirical analyses

The DNA methylation sites identified in the EWAS at P < 1×10^−7^ and the SNPs identified in the GWAS at P < 5×10^−8^ were mapped to genes and genesets. Overlap between genes was calculated as before (Equation (1)), enrichment scores were generated and correlated as described above.

Expected overlap was generated to compare to the observed results. For this, random positions were chosen in the genome equal to the number of DMPs identified in the EWAS. These genes were then used to assess gene and geneset overlap as for the observed results. This was repeated 1000 times to generate a null distribution and a z-test was used to assess whether there was a difference between the observed results and the mean of the null distribution.

There is a correlation structure within DNA methylation data (69), we hypothesised this might contribute to the observed results. By randomly sampling positions from the genome, a new correlation structure between DMPs would be generated. We tested whether sampling the genome in a non-random way, aimed at keeping some correlation structure, altered the results. To generate new data whilst attempting to keep a similar correlation structure, a fixed number of base pairs were added to each of the DMPs identified in the empirical analysis such that

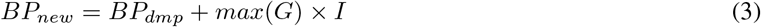

where *BP*_*new*_ = base pair of new site, *BP*_*dmp*_ = base pair of DMP identified in the EWAS, *G* = gene size, and *I* = iteration.

If *BP*_*new*_ extended beyond the end of a chromosome the position moved onto the next chromosome, with positions moving past the end of chromosome 22, being moved to chromosome 1.

Overall, the overlap between genes and genesets identified by EWAS and GWAS did not change across null distribution sampling methods.

### Understanding architecture from geneset overlap

Simulations were setup as illustrated in **Supplementary figure 4**. Here we describe simulations for a single trait. These were repeated for all traits. Firstly, SNPs identified in the GWAS and DMPs identified in the EWAS were mapped to genes as described above in Empirical analyses. Genes were then randomly sampled from Ensembl gene IDs and were assigned as either “causal,” meaning changes in that gene effect variation in the phenotype across individuals, or “associated,” meaning changes in the gene are associated with the phenotype across individuals, but the nature of association is not known. The empirically identified (or “known”) GWAS genes (*KGG*) were added to the list of causal genes and the empirically identified (or “known”) EWAS genes (*KEG*) were added to the list of associated genes. These combined set of causal and associated genes can be thought of as all the genes related to the trait of interest. A number of genes, equal to the number of *KGG* (*N*_*KGG*_), was sampled from the causal set of genes and assigned to be the “GWAS genes” in the simulations. A number of genes, equal to the number of *KEG* (*N*_*KEG*_), was sampled from the associated set of genes and assigned to be the “EWAS genes” in the simulations. Then geneset enrichment analyses for both the EWAS and GWAS genes were performed (equation (2)) and correlation between the enrichment scores was assessed as previously. In these simulations, the number of total genes was varied and the number of causal and associated genes was always set to be half of the total number of genes related to a trait. The total number of genes was proportional to the total number of known genes (*N*_*KT G*_ = *N*_*KGG*_ + *N*_*KEG*_). In each simulation, the number of associated genes (and causal genes) equalled *N*_*KT G*_ discovered multiplied by 1, 2, 3, 5, 10 or 20. Therefore, the smallest number of total genes for any simulation was double the number of *N*_*KT G*_ and the greatest number of total genes was 40 times *N*_*KT G*_. The other variable set to vary between simulations was the proportion of overlap between causal and associated genes. The proportion of overlap was 0, 0.01, 0.1, 0.5 or 1, where 0 represented the scenario where only the overlap in *KGG* and *KEG* would be present in the overlap between causal and associated genes and 1 represented the scenario where all causal and associated genes were the same. For each simulation scenario, the simulations were repeated 1000 times and box plots show the range of output from those 1000 repeats.

### Assessing the correlation between geneset enrichment results

GWAS were extracted from the IEU OpenGWAS Project (31,32) with the following criteria:

- Sample size > 5000
- European population
- For binary traits, number of cases and controls had to be greater than 500
- Full genome-wide results, i.e. not just associations between a molecular trait and variants in cis.

For each GWAS, all SNPs that associated with the trait at P<5×10^−8^ were extracted. CpGs associated with the 14 EWAS at P<1×10^−7^ were then extracted and mapped to genes. For each study, enrichment scores were generated for GO terms as before (Equation (2)) and correlation between them assessed.

When assessing whether gene overlap or geneset enrichment score correlations were greater than the mean, a z-test was performed. As multiple tests were performed we applied the Benjamini-Hochberg method (70) to limit the false discovery rate.

### Code availability

Code used to run the analyses is available here: https://github.com/thomasbattram/ewas-gwas-comparisons Analyses were completed using R (version 3.6.2) and Python (version 3.7.4).

## Data Availability

All data are available at http://www.ewascatalog.org/,  https://gwas.mrcieu.ac.uk/ and http://www.epigraphdb.org/

https://gwas.mrcieu.ac.uk/

http://www.epigraphdb.org/

## Acknowledgements

We are grateful to Dr. Kaitlin Wade for highly valuable comments.

