## Supplementary Material for "A comparison of the genes and genesets identified by EWAS and GWAS of fourteen complex traits"

### Figures

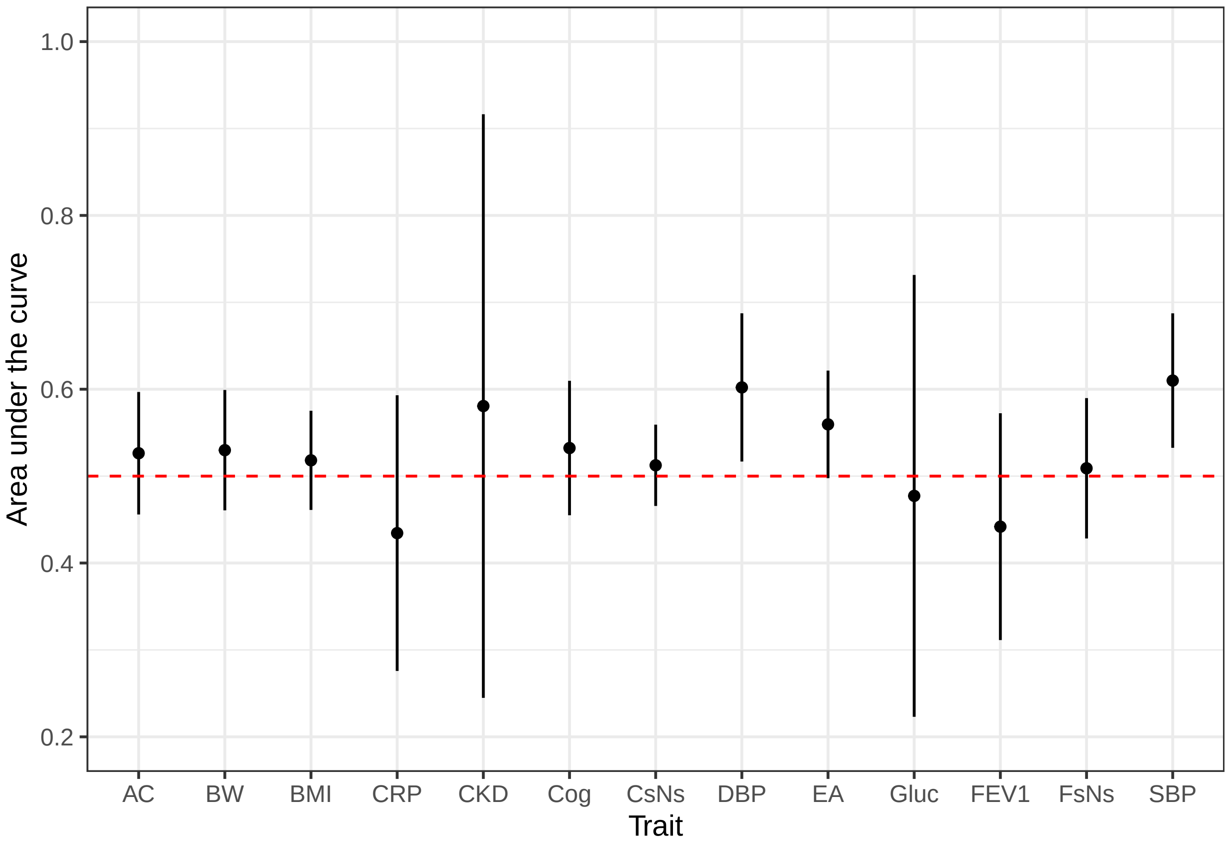


Figure 1: **Can genetic variant associations predict the presence of DNA methylation associations in the same region?.** For each 500kb region in the genome, the largest SNP-trait effect size was extracted. ROC curves were produced to determine whether these could predict whether a differentially methylated position related to the same trait was present in the same 500kb region. The area under these curves (AUC), with their confidence intervals, are plotted for each trait. The red dashed line is at AUC = 0.5, which represents a prediction no better than chance. AC = alcohol consumption per day, BW = birthweight, BMI = body mass index, CKD = chronic kidney disease, Cog = cognitive ability (digit test), CRP = c-reactive protein, CsNs = current smokers vs never smokers, DBP = diastolic blood pressure, EA = educational attainment, Gluc = fasting glucose, FEV1 = forced expiratory volume in one second, FsNs = former smokers vs never smokers, SBP = systolic blood pressure.

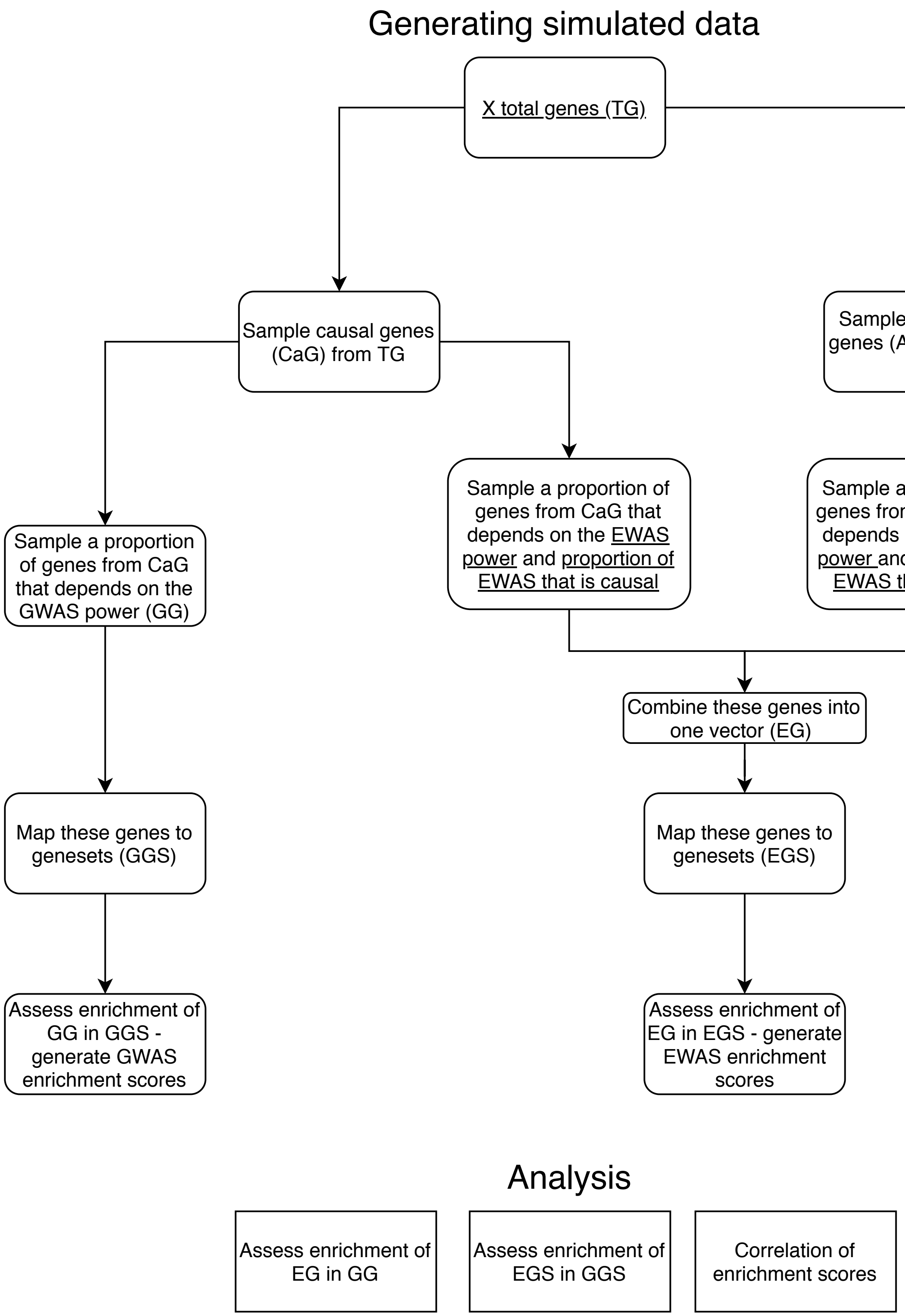


Figure 2: **Flowchart demonstrating how simulations were set up.** The data was simulated under the flowchart specified under "Generating simulated data" and then data was analysed as specified under "Analysis." Underlined variables were varied. The simulations were repeated 1000 times for each set of parameters.

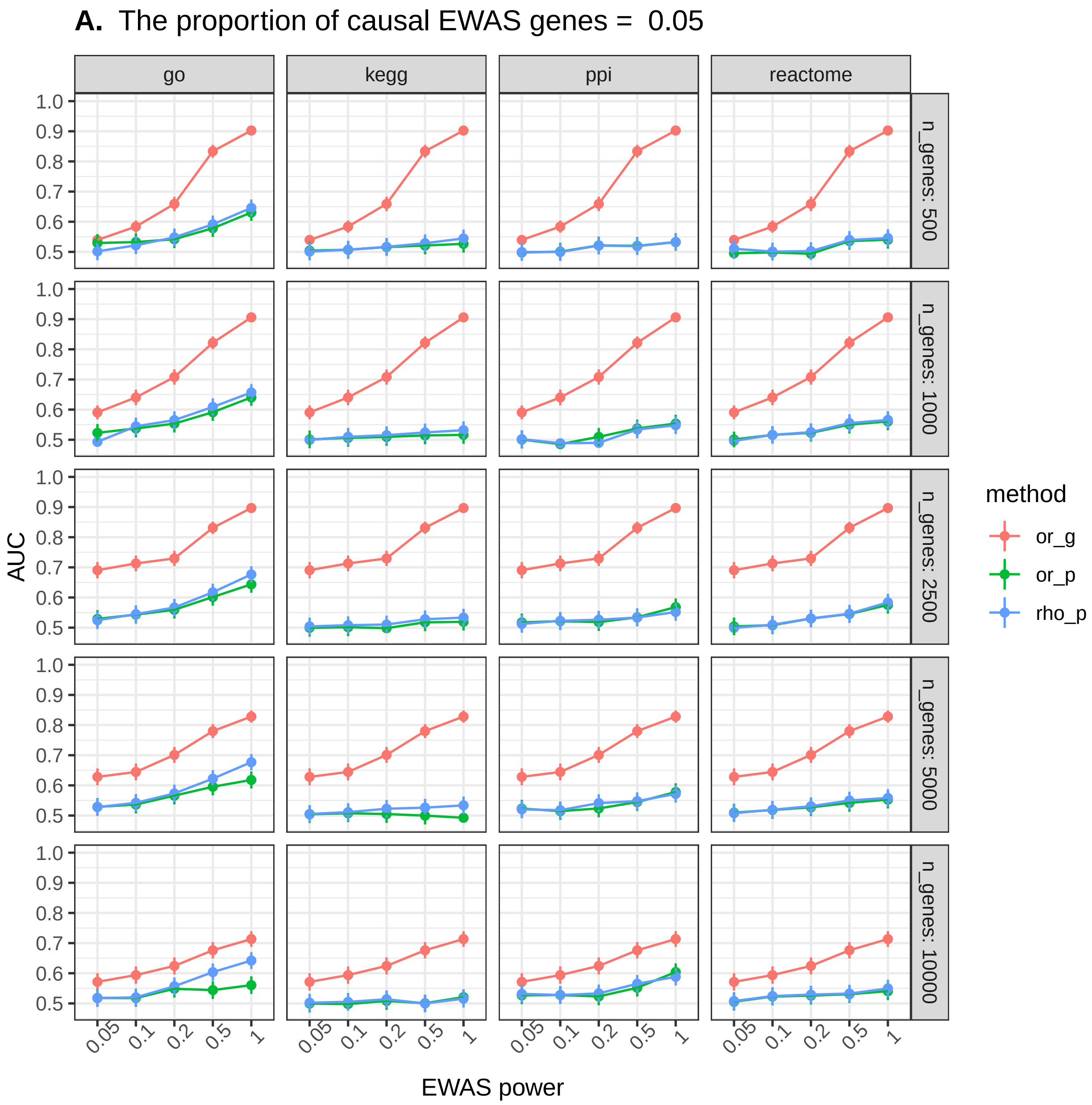
A. The proportion of causal EWAS genes = 0.05
go
kegg
ppi
reactome
n_genes: 500
n_genes: 1000
n_genes: 2500
n_genes: 5000
n_genes: 10000
AUC
1.0
0.9
0.8
0.7
0.6
0.5
0.05
0.1
0.2
0.5
1
EWAS power
method
or_g
or_p
rho_p

6

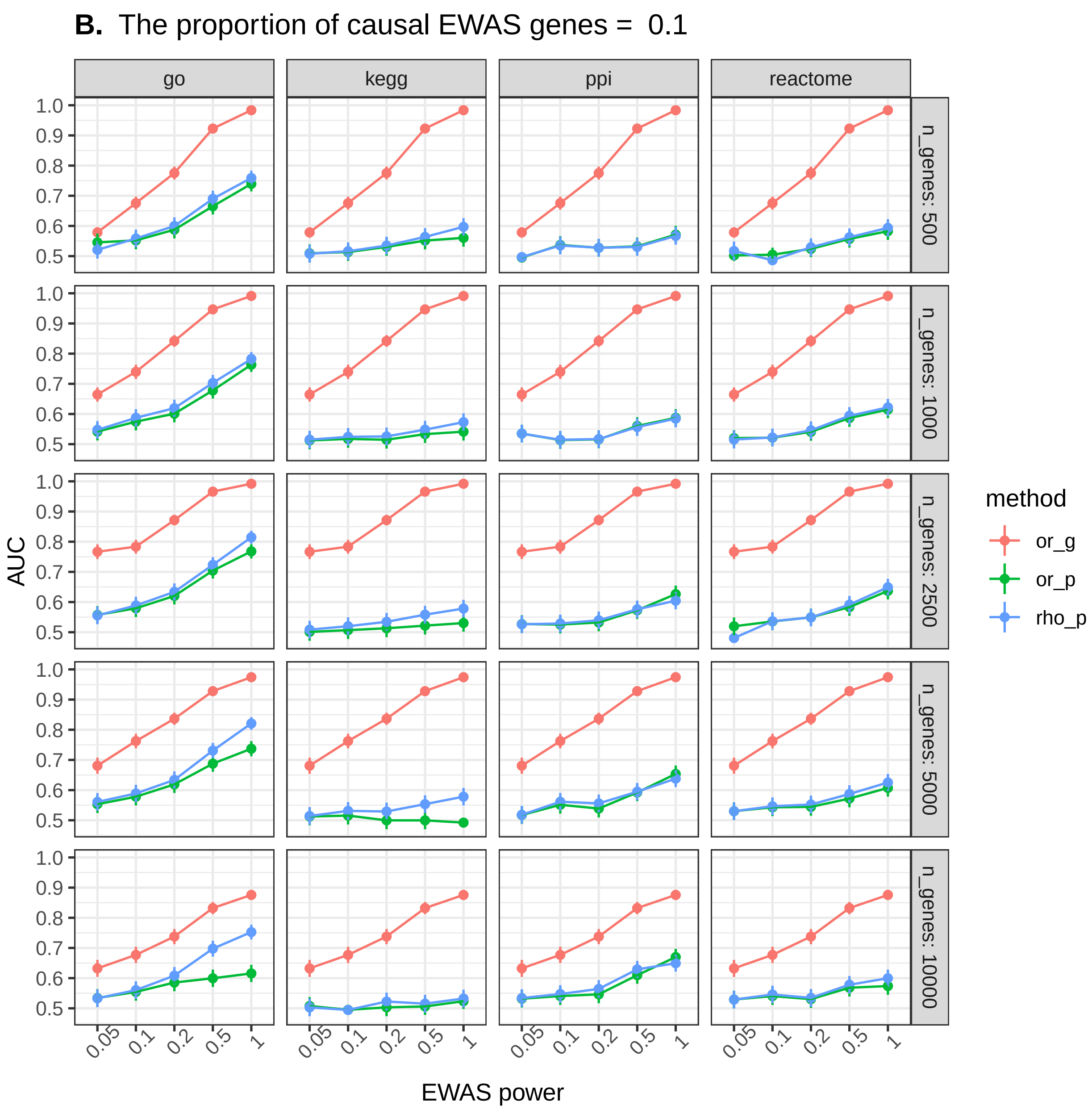

B. The proportion of causal EWAS genes = 0.1
go
kegg
ppi
reactome
n_genes: 500
n_genes: 1000
n_genes: 2500
n_genes: 5000
n_genes: 10000
AUC
1.0
0.9
0.8
0.7
0.6
0.5
0.05
0.1
0.2
0.5
1
EWAS power
method
or_g
or_p
rho_p


7

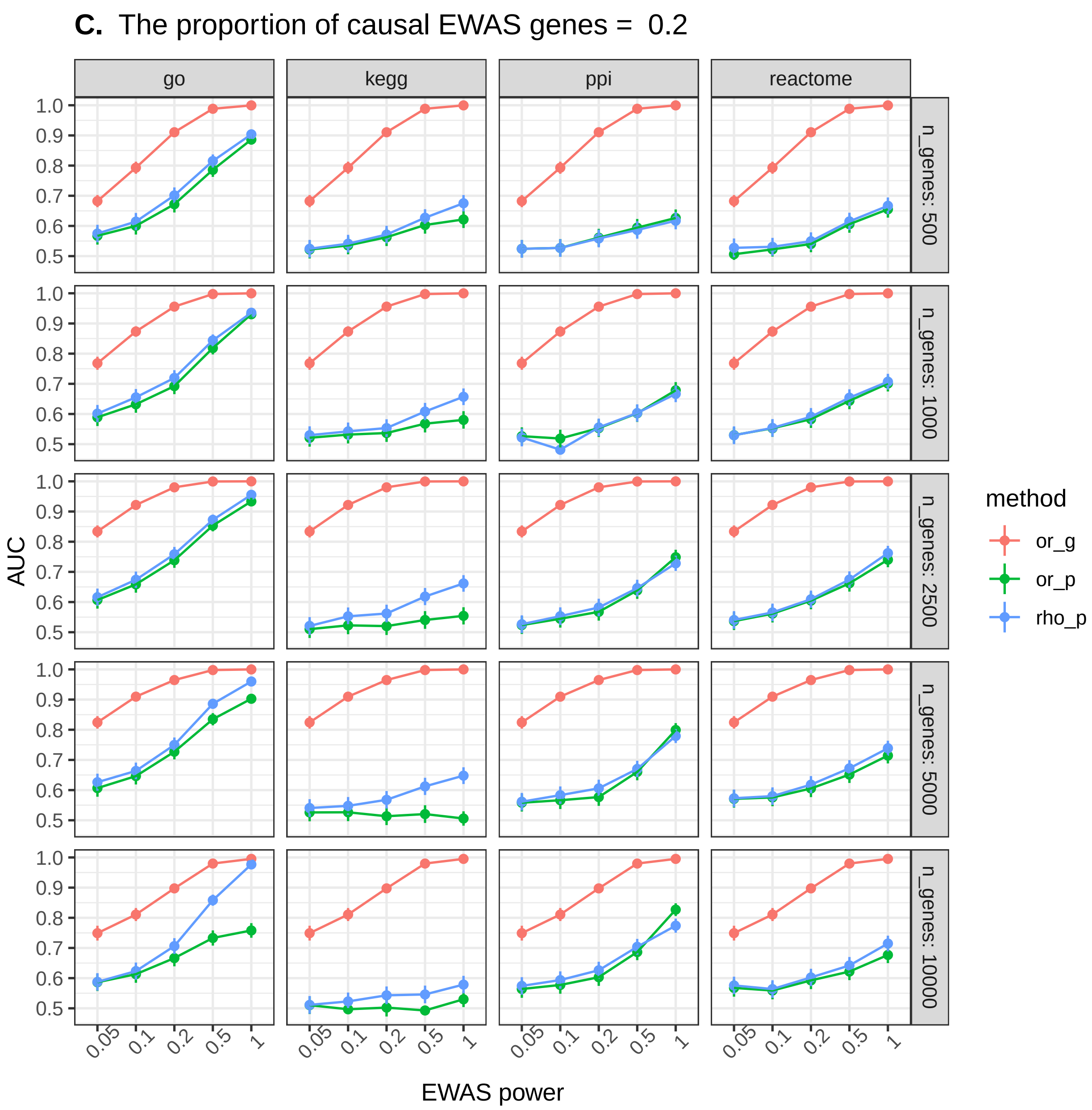
C. The proportion of causal EWAS genes = 0.2
go
kegg
ppi
reactome
n_genes: 500
n_genes: 1000
n_genes: 2500
n_genes: 5000
n_genes: 10000
AUC
EWAS power
method
or_g
or_p
rho_p

8

**D.** The proportion of causal EWAS genes = 0.5

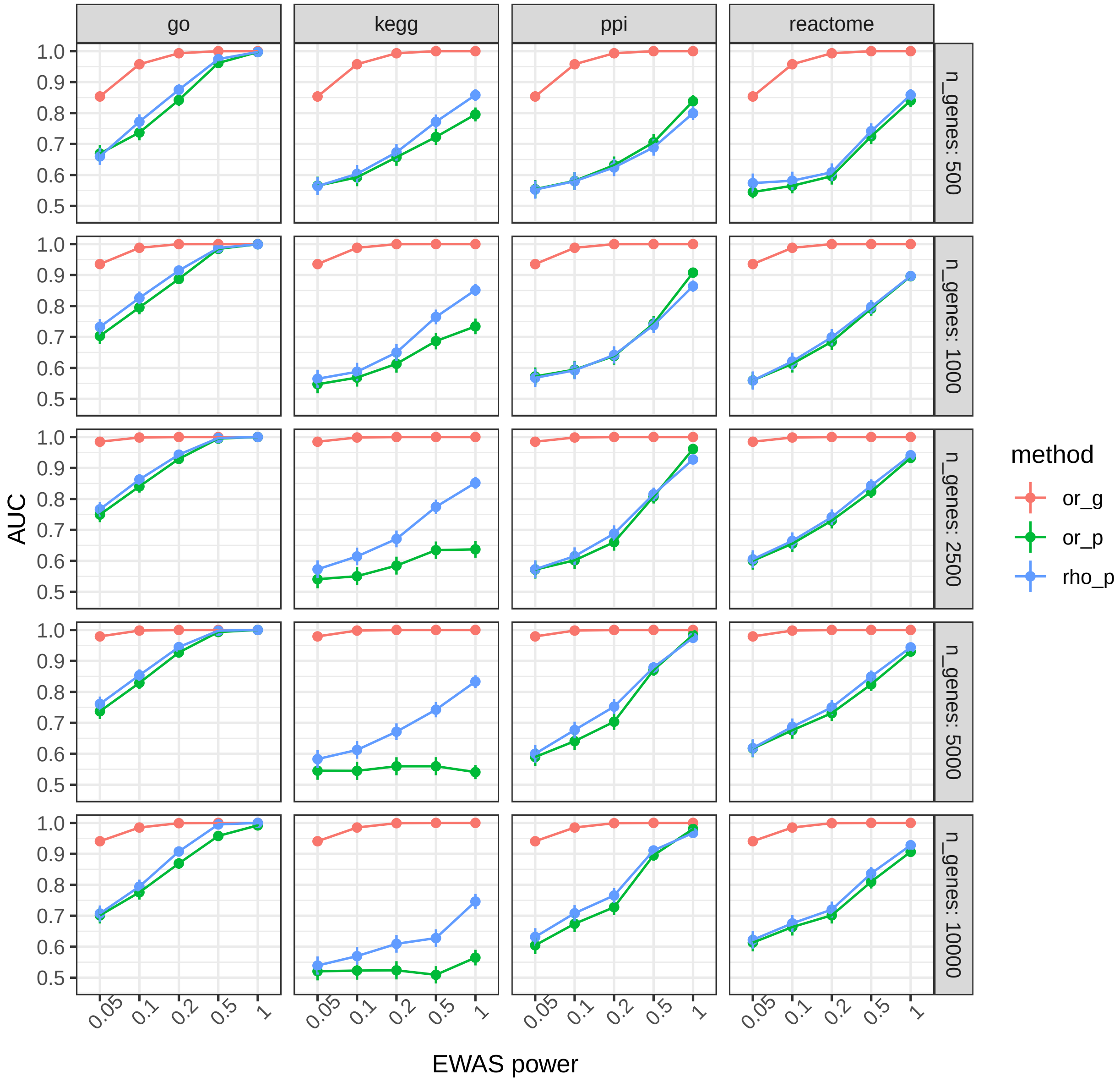


9

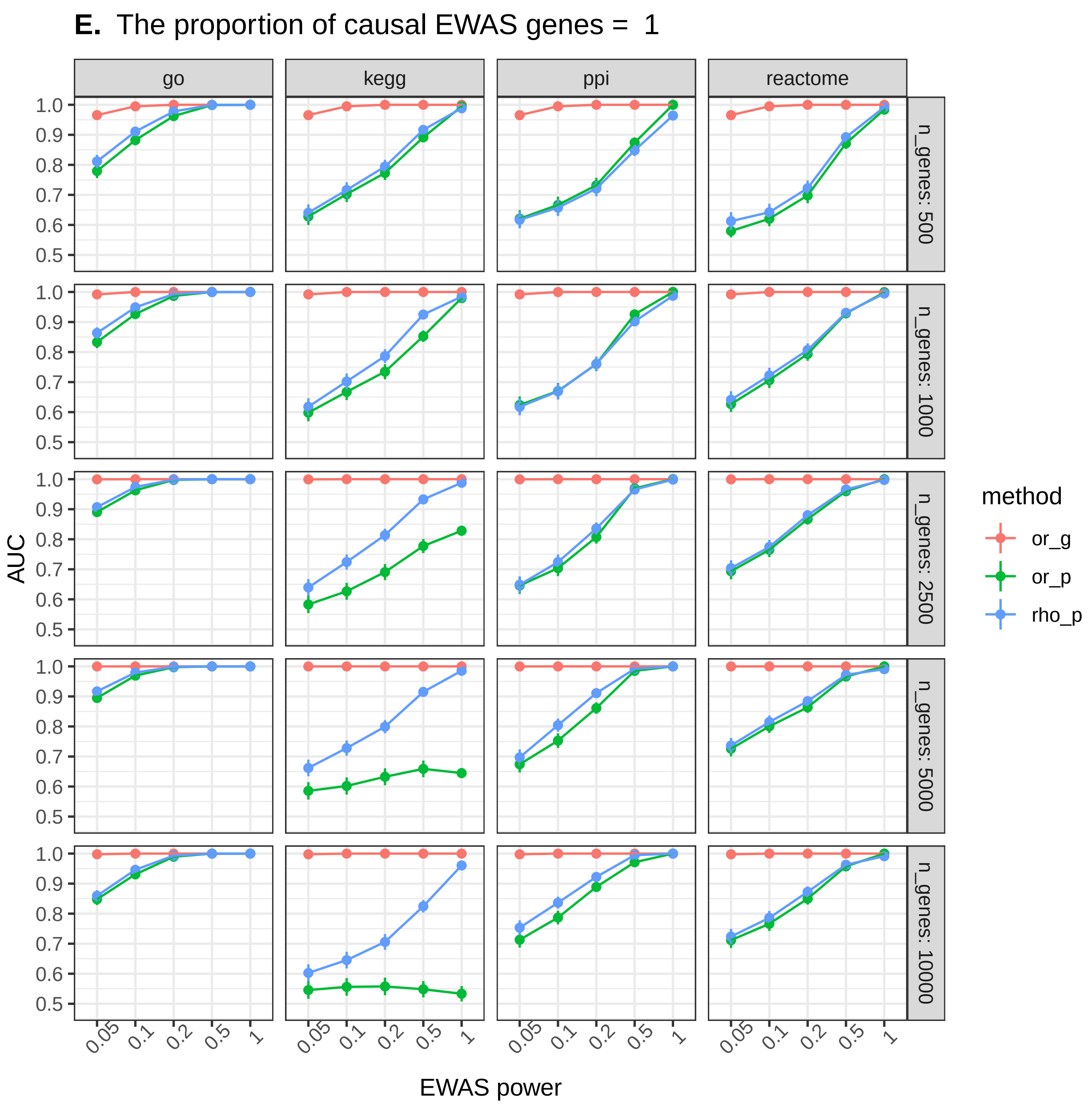


Figure 3: **Power to detect overlap between genes and genesets identified by corresponding EWAS and GWAS.** Simulations were set up as illustrated in **Supplementary figure 2**. The ability to distinguish between results generated when EWAS and GWAS were sampling, in part, from the same set of causal genes and results generated when EWAS was sampling random genes from the genome. The header of each set indicates the proportion of genes identified by the simulated EWAS that were set to be causal. or_g = assessing overlap of genes, or_p = assessing overlap of genesets, rho_p = assessing correlation between geneset enrichment scores. go = gene ontology, ppi = protein-protein interaction database from EpiGraphDB.

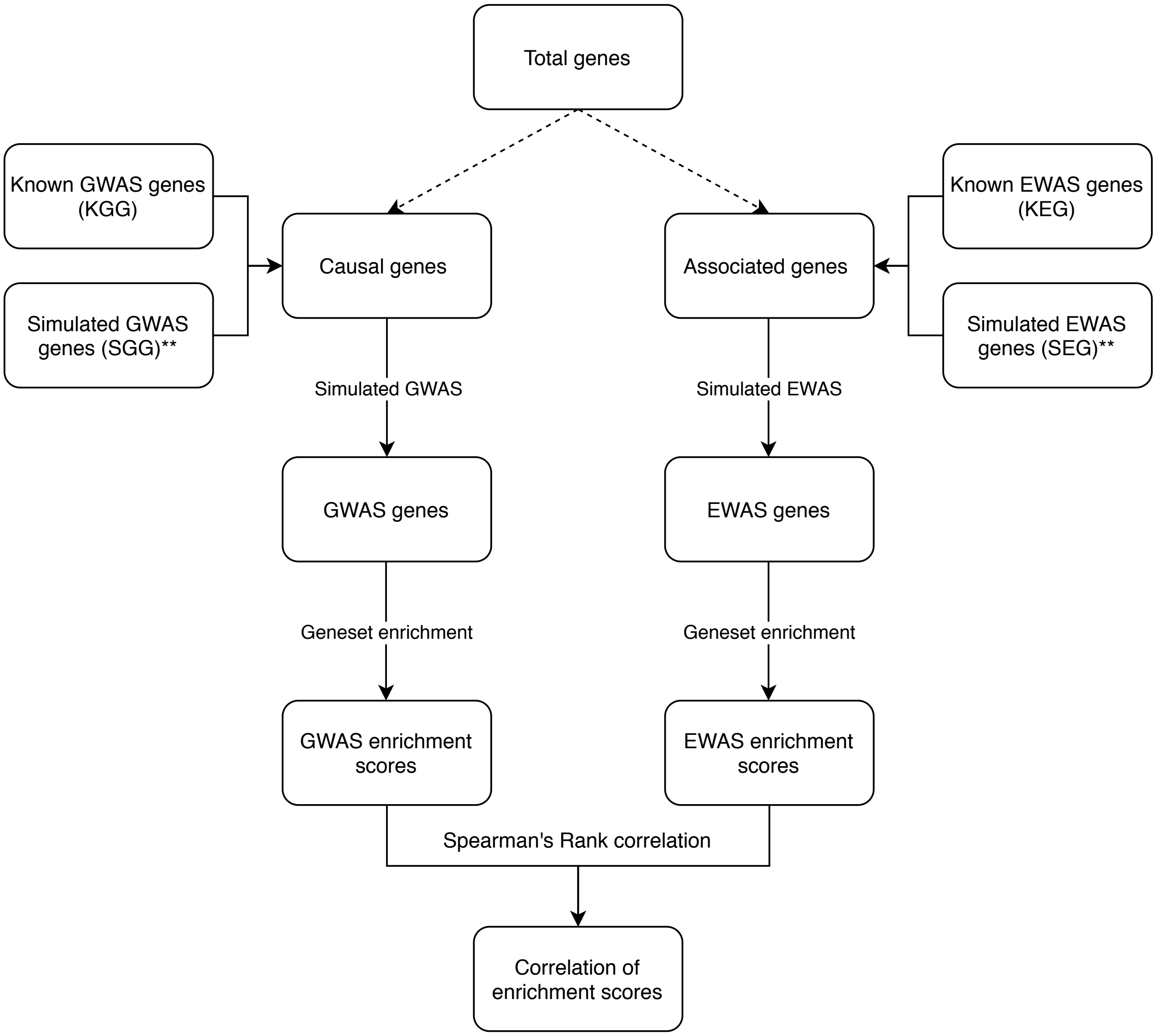


Figure 4: **Flowchart demonstrating how the second set of simulations were set up for each trait.** Phenotypic variation will be caused by changes in gene/protein polymers (causal genes) and can be associated with changes in gene/protein polymers via other routes such as confounding or reverse causation (associated genes). In these simulations the causal genes were a mix of genes identified by GWAS of that trait, known GWAS genes (KGG), and a randomly selected set of genes, simulated GWAS genes (SGG). The associated genes were a mix of genes identified by EWAS of that trait, known EWAS genes (KEG), and a randomly selected set of genes, simulated EWAS genes (SEG). The level of overlap in the causal and associated genes was modified by changing the overlap in the SGG and SEG. The number of causal and associated genes was kept the same for each simulation, but this number varied between simulations. The minimum number of causal genes and the minimum number of associated genes was equal to the sum of KGG and KEG. The "simulated GWAS" step in the simulation simply equates to randomly sampling from the causal genes. The number of genes sampled was equal to the number of KGG. The "simulated EWAS" step was identical except the number of KEG from the associated genes. Geneset enrichment was performed as described in the **Methods**. The simulations were repeated 1000 times for each set of parameters.

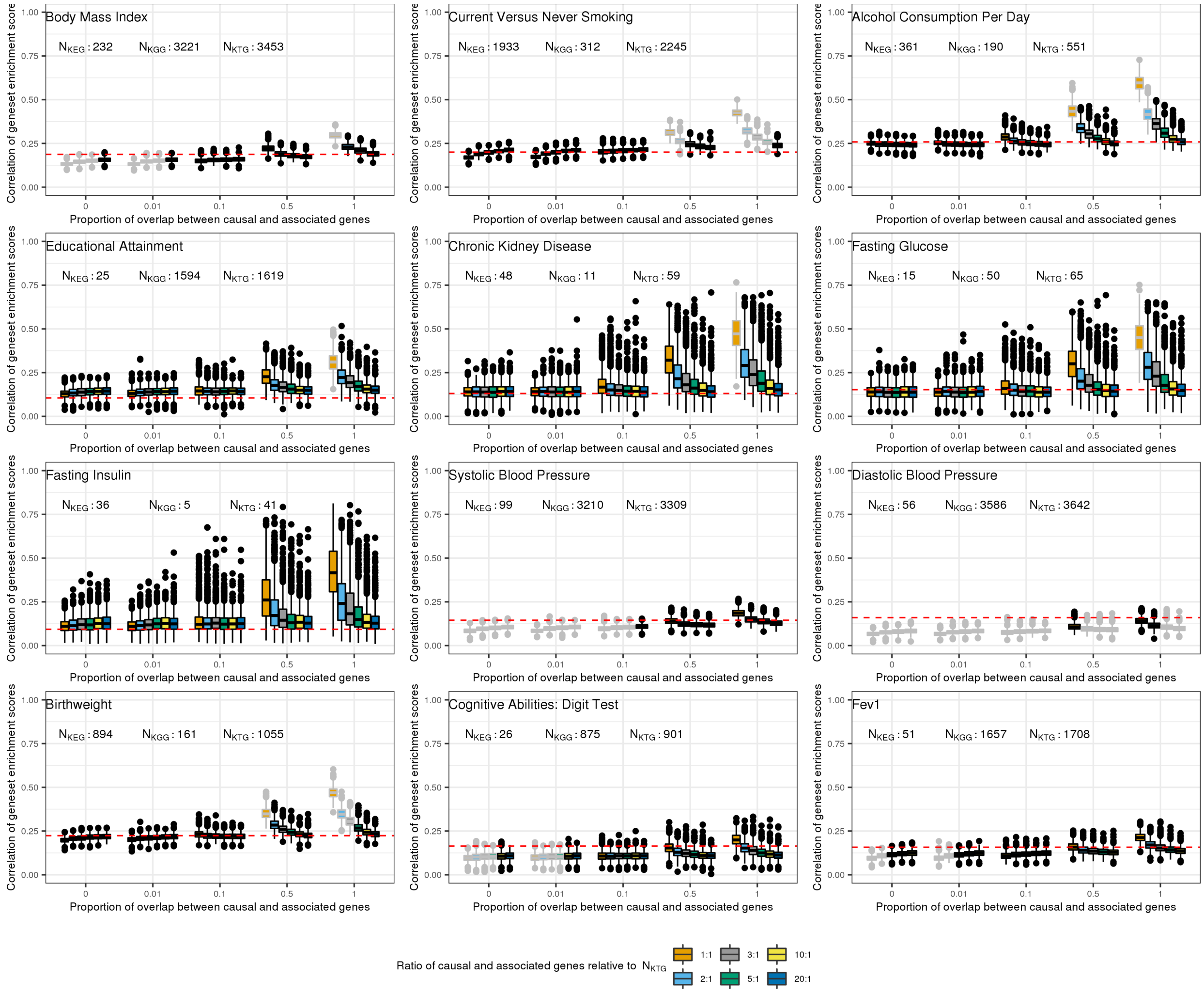


Figure 5: **Simulations to understand the likely number of genes still to identify in EWAS and GWAS of five traits under different trait architectures**. Simulations were set up as illustrated in **Supplementary figure 4**. Correlation of geneset enrichment scores from empirical data (**Table 3**), is shown as a red dashed line. Box plots show the range of enrichment score correlations from 1000 simulations using the parameters indicated. The number of causal and associated genes, as well as the overlap between these genes were varied. $N_{KEG}$ = number of known EWAS genes, $N_{KEG}$ = number of known GWAS genes, $N_{KTG}$ = number of known total genes ($N_{KEG}$ + $N_{KGG}$). By way of an example, when $N_{KTG}$ = 557 and the ratio of causal and associated genes relative to $N_{KTG}$ is 1:1, the number of causal genes in the simulations will be 557 and the number of associated genes in the simulations will be 557. Scenarios which lie close to the empirical result (red dashed line) are more likely to reflect the true underlying number of genes related to a trait and the true overlap between the causal and associated genes.

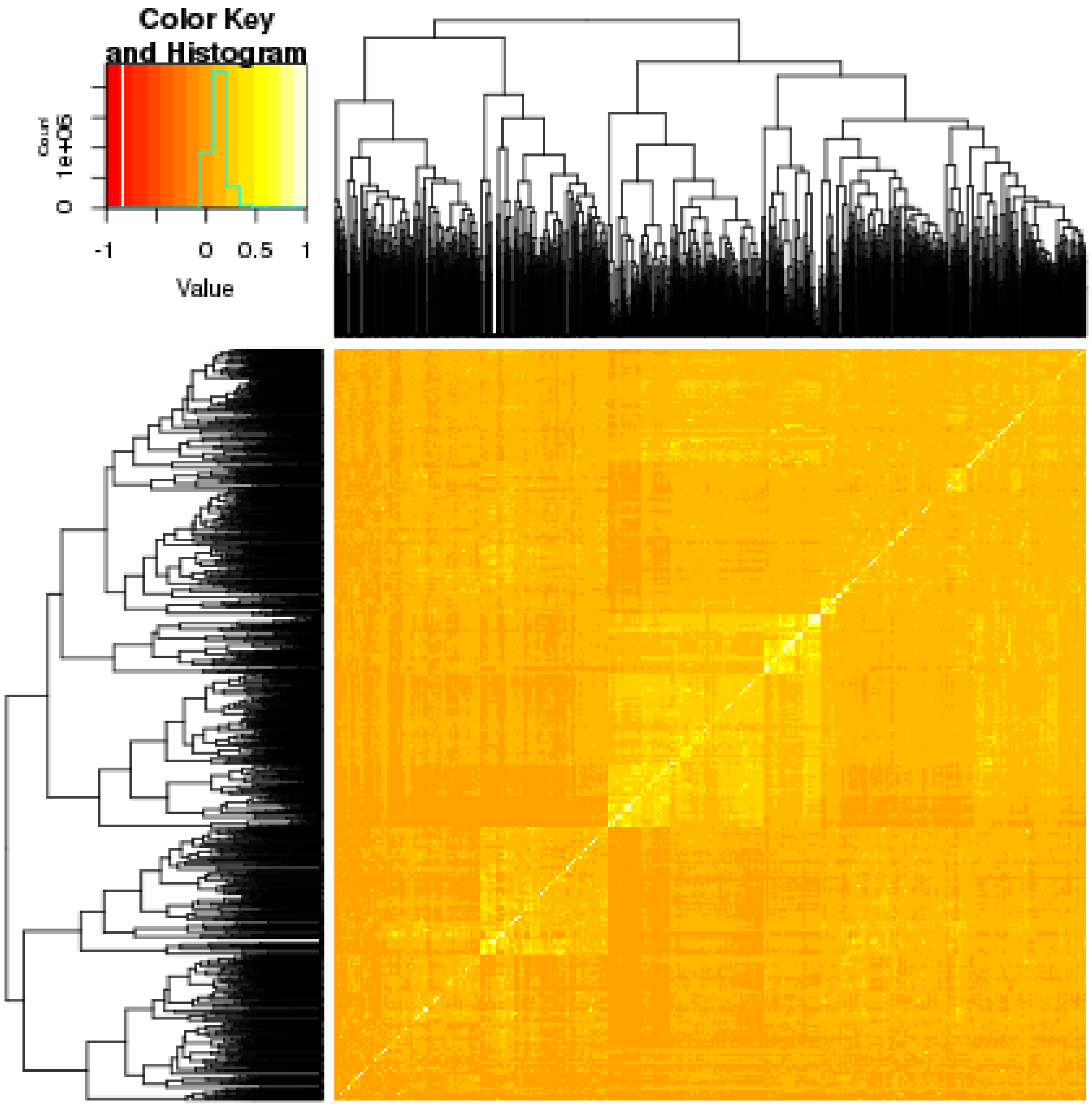


Figure 6: **Correlation across geneset enrichment scores for 1886 GWAS and 14 EWAS.**

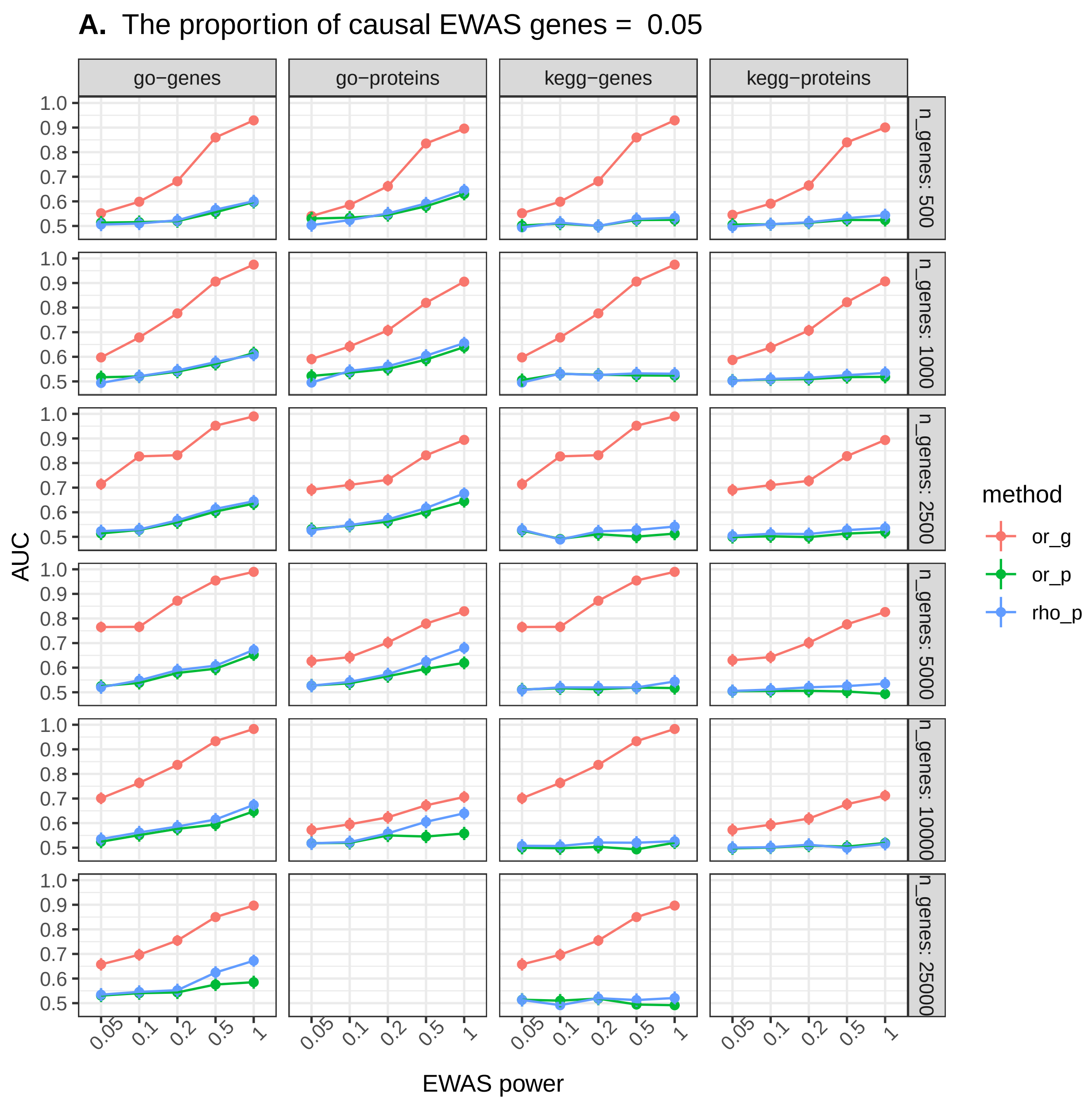
A. The proportion of causal EWAS genes = 0.05
go−genes
go−proteins
kegg−genes
kegg−proteins
n_genes: 500
n_genes: 1000
n_genes: 2500
n_genes: 5000
n_genes: 10000
n_genes: 25000
AUC
EWAS power
0.05
0.1
0.2
0.5
1
method
or_g
or_p
rho_p

**B.** The proportion of causal EWAS genes = 0.1

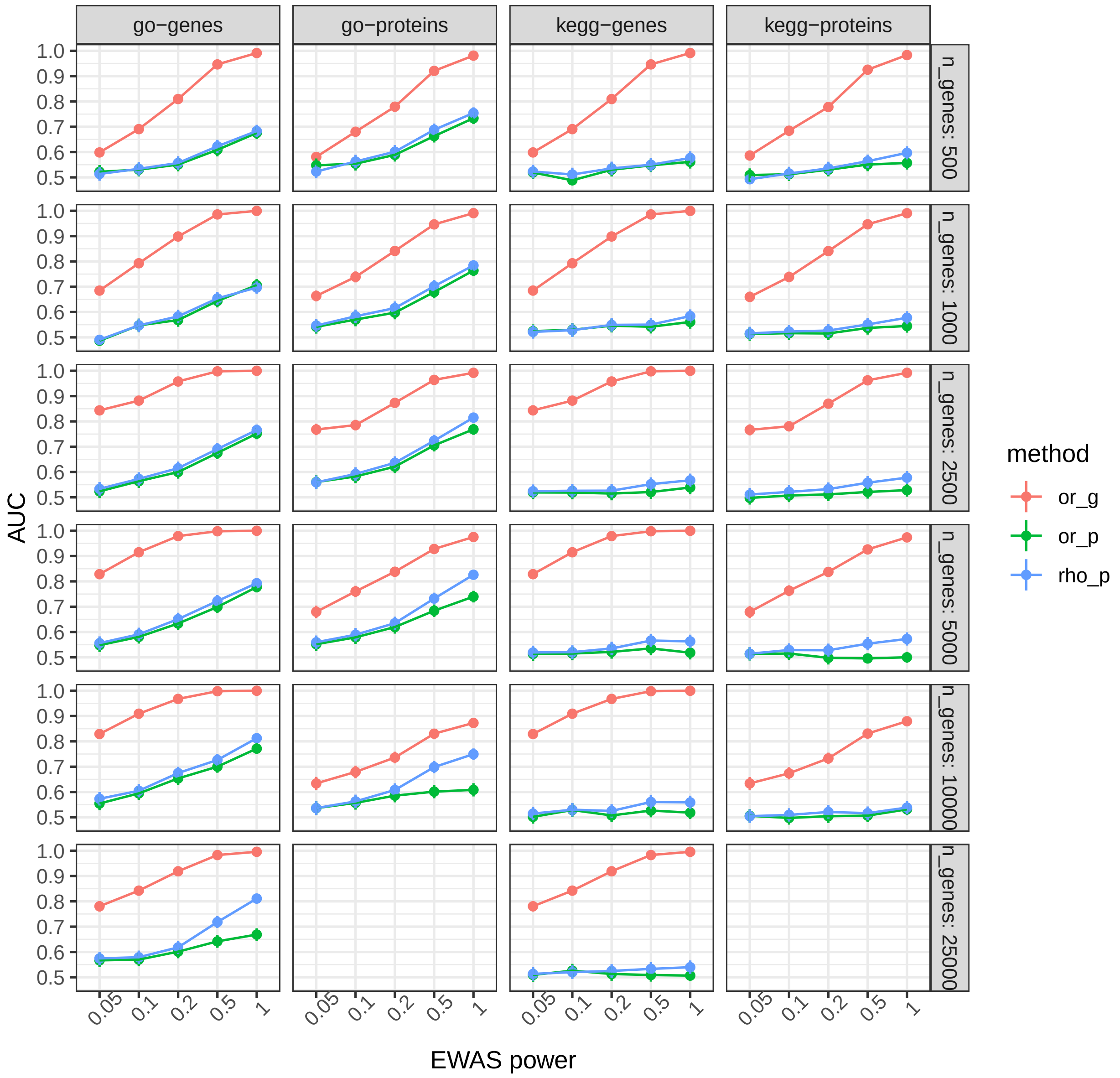


11

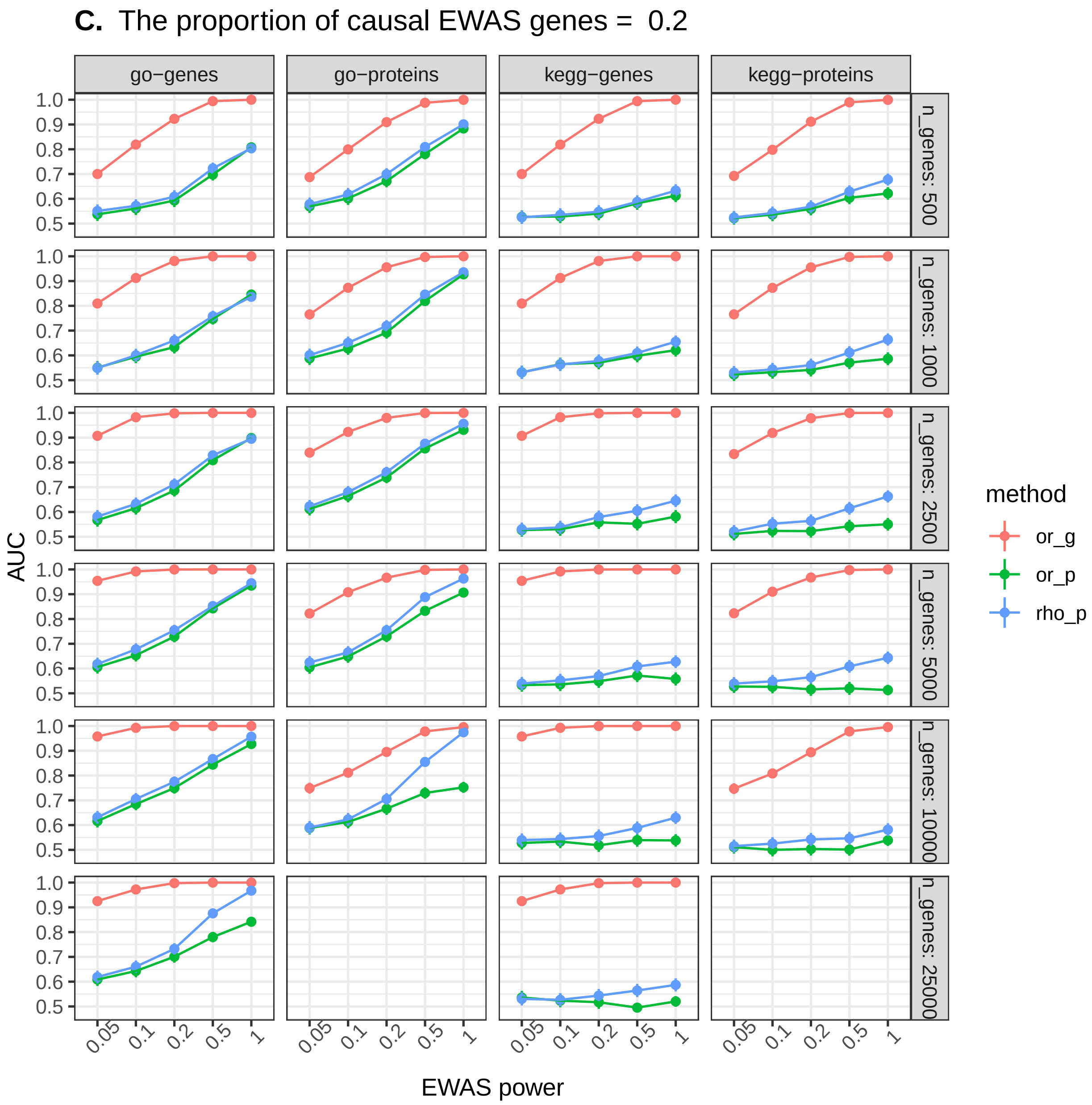
C. The proportion of causal EWAS genes = 0.2
go−genes
go−proteins
kegg−genes
kegg−proteins
n_genes: 500
n_genes: 1000
n_genes: 2500
n_genes: 5000
n_genes: 10000
n_genes: 25000
AUC
1.0
0.9
0.8
0.7
0.6
0.5
0.05
0.1
0.2
0.5
1
EWAS power
method
or_g
or_p
rho_p

12

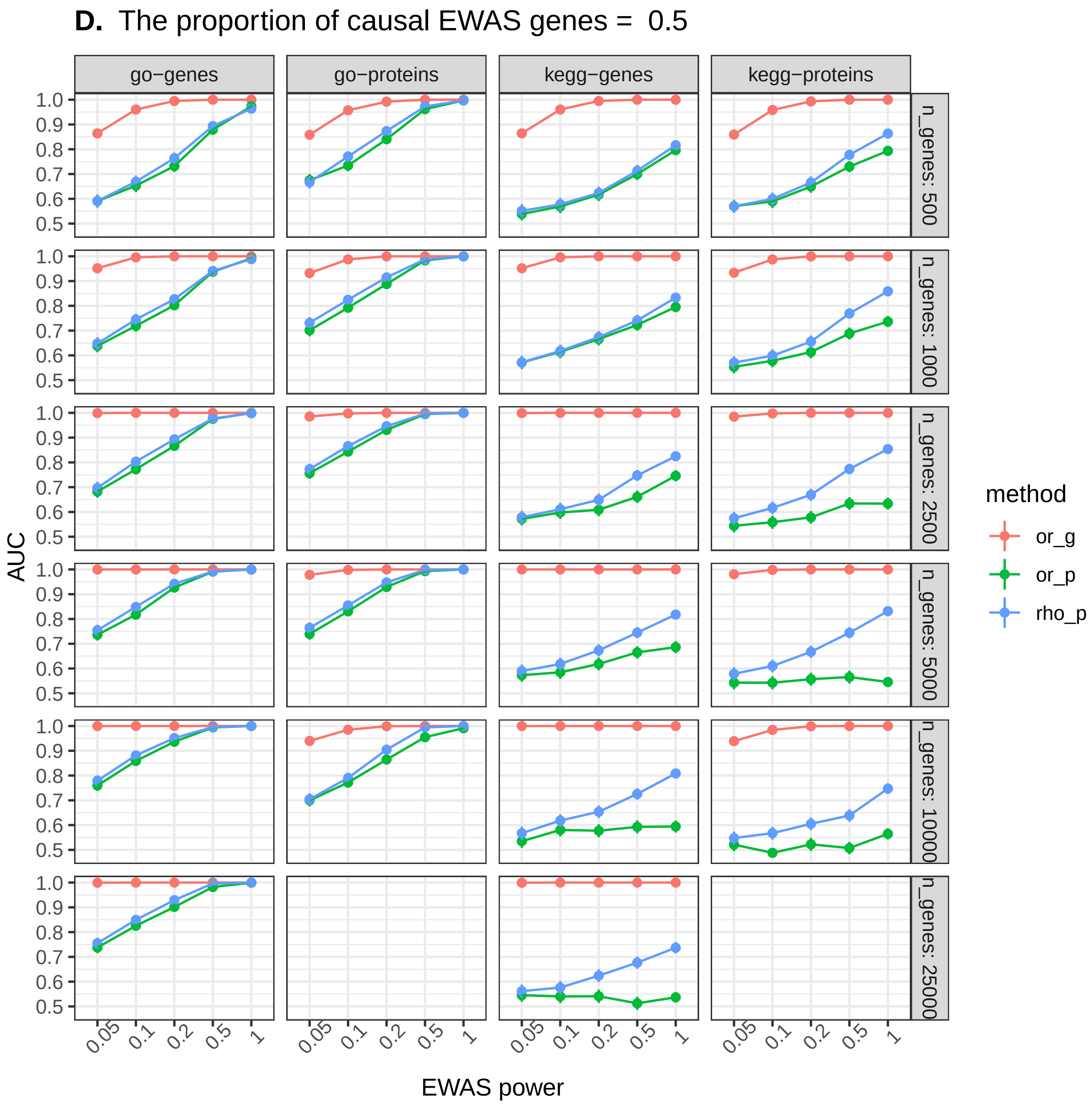

D. The proportion of causal EWAS genes = 0.5
go−genes
go−proteins
kegg−genes
kegg−proteins
n_genes: 500
n_genes: 1000
n_genes: 2500
n_genes: 5000
n_genes: 10000
n_genes: 25000
AUC
EWAS power
0.05
0.1
0.2
0.5
1
0.5
0.6
0.7
0.8
0.9
1.0
method
or_g
or_p
rho_p

**E.** The proportion of causal EWAS genes = 1

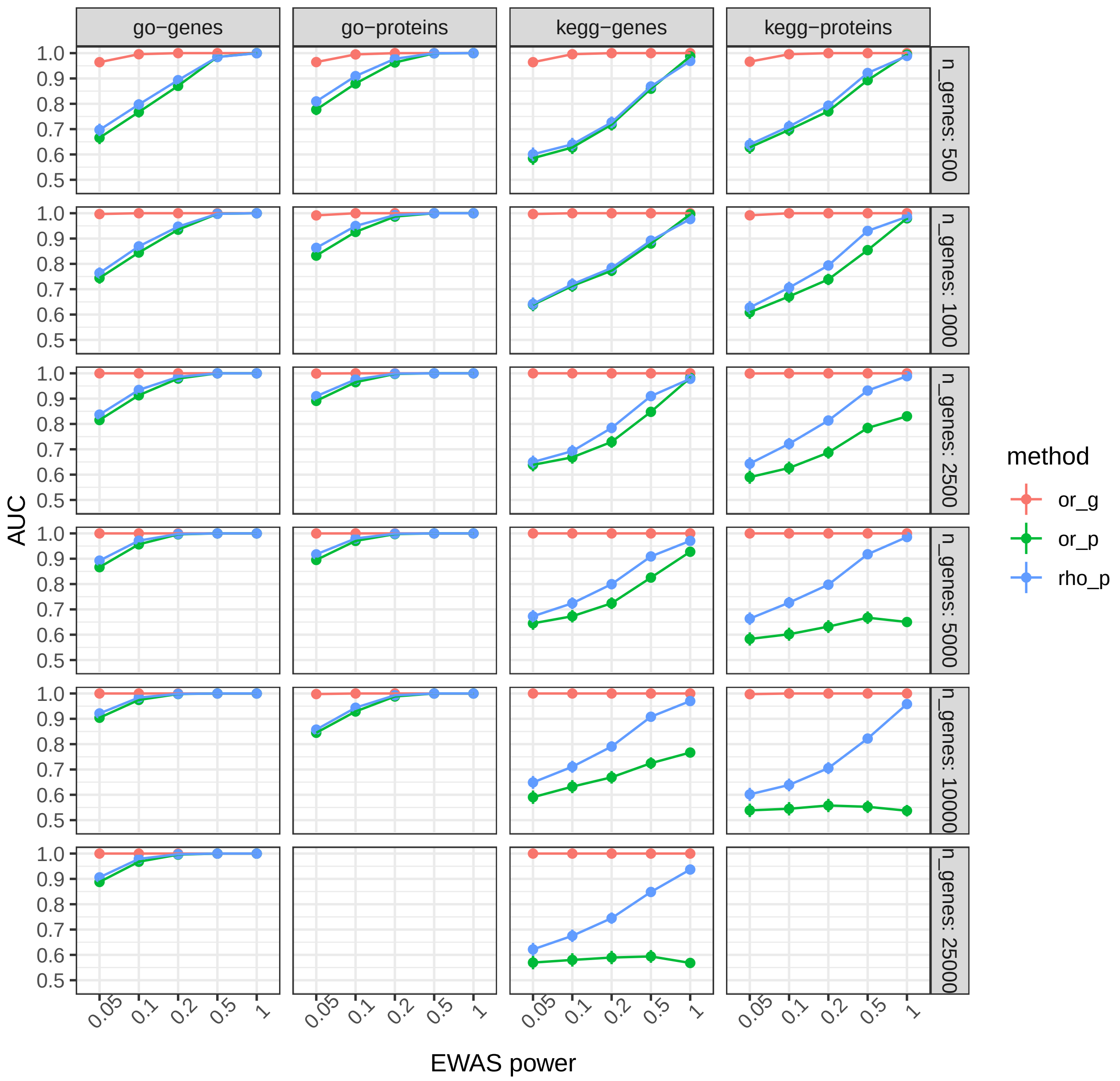


Figure 7: **Power to detect overlap between genes and genesets identified by corresponding EWAS and GWAS when mapping signal to all genes and protein coding genes.** Simulations were set up as illustrated in **Supplementary figure 2**. The ability to distinguish between results generated when EWAS and GWAS were sampling, in part, from the same set of causal genes and results generated when EWAS was sampling random genes from the genome. The header of each set indicates the proportion of genes identified by the simulated EWAS that were set to be causal. or_g = assessing overlap of genes, or_p = assessing overlap of genesets, rho_p = assessing correlation between geneset enrichment scores. go = gene ontology, suffix of '-genes' denotes using all Ensembl gene IDs for the analysis and the suffix of '-proteins' denotes using only protein coding genes.
